# The unmitigated profile of COVID-19 infectiousness

**DOI:** 10.1101/2021.11.17.21266051

**Authors:** Ron Sender, Yinon M. Bar-On, Sang Woo Park, Elad Noor, Jonathan Dushoff, Ron Milo

**Author notes:** Equal contribution (Ron Sender, Yinon M. Bar-On).

## Abstract

Quantifying the temporal dynamics of infectiousness of individuals infected with SARS-CoV-2 is crucial for understanding the spread of the COVID-19 pandemic and for analyzing the effectiveness of different mitigation strategies. Many studies have tried to use data from the onset of symptoms of infector-infectee pairs to estimate the infectiousness profile of SARS-CoV-2. However, both statistical and epidemiological biases in the data could lead to an underestimation of the duration of infectiousness. We correct for these biases by curating data from the initial outbreak of the pandemic in China (when mitigation steps were still minimal), and find that the infectiousness profile is wider than previously thought. For example, our estimate for the proportion of transmissions occurring 14 days or more after infection is an order of magnitude higher - namely 19% (95% CI 10%-25%). The inferred generation interval distribution is sensitive to the definition of the period of unmitigated transmission, but estimates that rely on later periods are less reliable due to intervention effects. Nonetheless, the results are robust to other factors such as the model, the assumed growth rate and possible bias of the dataset. Knowing the unmitigated infectiousness profile of infected individuals affects estimates of the effectiveness of self-isolation and quarantine of contacts. The framework presented here can help design better quarantine policies in early stages of future epidemics using data from the initial stages of transmission.

## Introduction

In an emerging epidemic, such as the current COVID-19 pandemic, information about key epidemiological parameters of the causative infectious agent (SARS-CoV-2 in the case of COVID-19) is crucial for monitoring and mitigating the spread of the disease. A central epidemiological parameter which determines the time scale of transmission is the generation interval - the time between the infection of the infector (first case) and of the infectee (secondary case). Measuring the generation interval directly is hard in practice, as determining the exact time of infection is challenging. Thus, to infer the generation interval for an emerging infectious disease, researchers usually rely on two widely reported epidemiological parameters: the *incubation period* - the time between infection with the virus and the onset of symptoms (either for the infector or the infectee) - and the *serial interval -* the time between onset of symptoms of the infector and infectee ^1,2^ (Figure 1). Key epidemiological delays, such as incubation periods, serial intervals, and generation intervals, vary across hosts and transmission events, and are thus described as distributions rather than fixed values.

**Figure 1:**
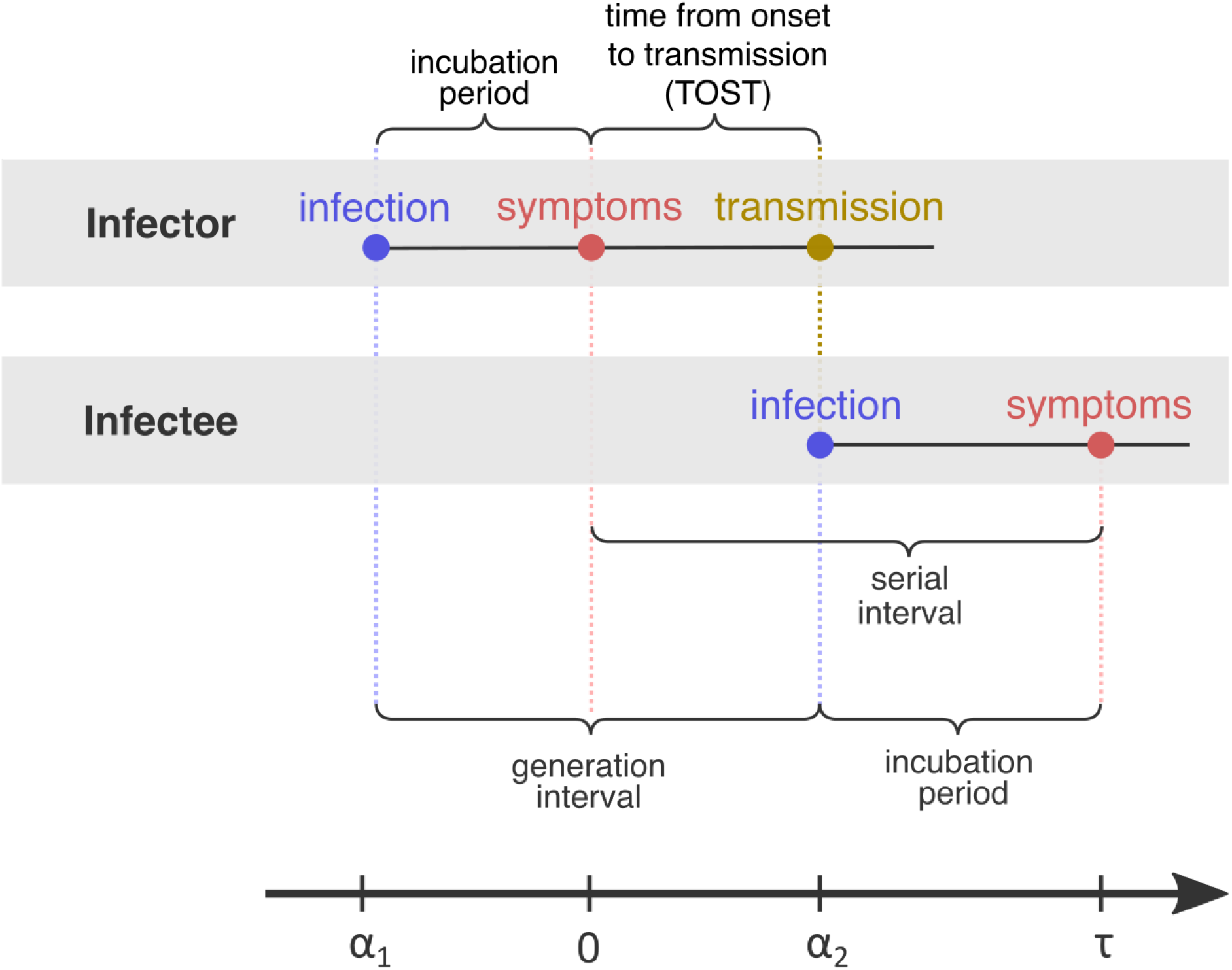
Definitions of epidemiological time intervals. The incubation period is defined as the time between infection and symptom onset (=− α_1_ for the infector, τ – α_2_ for the infectee). The serial interval (=τ) is defined as the interval between the onset of symptoms of two subsequent transmission events (infector and infectee) and the generation interval is the time lapse between the infections of those individuals (=α_2_ – α_1_). TOST stands for Time from Onset of Symptoms to Transmission ^3^, and is defined accordingly as the time lapse between symptom onset in the infector and the infection of the infectee (i.., transmission time). The timeline at the bottom corresponds to the notation used in the Methods section.

The generation-interval distribution plays a key role in determining the spread and control of emerging epidemics such as the ongoing COVID-19 pandemic. At the population-level, the generation-interval distribution links incidence of infection, particularly the epidemic growth rate r, with the reproduction number R ^4,5^. At the individual level, it characterizes the infectiousness profile (i.e., the temporal evolution of infectiousness from the time of infection). In the case of COVID-19, short generation intervals, driven by presymptomatic transmission, have limited the effectiveness of different mitigation strategies, including contact tracing ^6^, case isolation, quarantine ^7^, and testing ^8,9^.

The generation- and serial-interval distributions can change over the course of an epidemic. For example, they are affected by the behavior of the population and can be shortened by the introduction of mitigation steps such as social distancing and case isolation, which limit the spread of the disease and reduce the probabilities of transmission after symptom onset ^10^. Our study aims to estimate the temporal dynamics of transmissibility of infected cases in the absence of intervention measures, noted hereafter as the “unmitigated generation interval”. Unbiased estimates of the time profile of transmissibility are important for inferring the effectiveness of self-isolation or quarantine policies in the absence of other interventions.

In practice, estimating the unmitigated infectious profile is expected to be challenging, since even in the absence of any mitigation policies, symptomatic individuals may self-isolate, reducing their own chances of late transmission. To address this issue, we apply a strict data curation procedure to account for which transmission events occurred both before major mitigation steps took place and before awareness of the epidemic became widespread. Most available estimates of the generation-interval distribution addressed the effects of mitigation only in a limited manner, not fully accounting for steps such as contact tracing and case isolation ^3,11,12^

Even after minimizing mitigation and behavioral effects, estimating the generation-interval distribution directly from contact tracing data remains difficult because the time-point of infection of both the infector and the infectee are usually unknown. Instead, researchers estimate generation-interval and incubation-period distributions by calculating the likelihood of observing all serial intervals in the transmission pair dataset ^3,6,11^ (or else, they simply use the serial-interval distribution as a proxy for the generation-interval distribution ^13^).

While the serial-interval-based framework has been widely applied to infer the generation-interval distribution of COVID-19 ^3,6,7,11,12^, there are several key methodological issues that could lead to considerable biases. First, the distribution of realized serial intervals depends on the rate of the spread of the disease as well as the direction from which they are measured: either forward from a cohort of infectors who developed symptoms at the same time, or backward from a cohort of infectees ^14^. For example, when the incidence of infection is increasing exponentially, individuals are more likely to have been infected recently. Therefore, a cohort of infectors that developed symptoms at the same time will have shorter incubation periods than their infectees on average, which will in turn affect the shape of the forward serial-interval distribution. Instead, most analyses of serial-interval distributions assume that the incubation periods of the infector and infectee follow the same distribution ^7,11,12^, and only a few studies partially account for this dynamical bias ^3,6^. Second, incubation periods and temporal profile of infectiousness are likely to be correlated across infectors - i.e, individuals that show symptoms later or earlier are also more likely to infect others later or earlier, respectively. Most available studies make strict assumptions on the relationship between the incubation period and the generation interval - either assuming that they are independent ^6,7,12^ or that the time from onset of symptoms to transmission (TOST) is independent of the incubation period ^11^. Only a few studies have compared various correlation models ^3^ or explicitly modeled the infectiousness profile relative to the incubation period ^15^. Finally, biases can arise from the data collection process. For example, determining who infected whom based on their symptom-onset dates can miss presymptomatic transmission. Likewise, long serial intervals may represent multiple chains of transmissions where intermediate hosts were not correctly identified. These biases can cause overestimation of the mean serial interval as well as the mean generation interval.

Currently, no available estimate for the generation interval deals with all the biases described above, impairing our ability to accurately describe the infectiousness of SARS-CoV-2-infected individuals in the absence of interventions. Here, we aggregate all available transmission data for Wuhan, China, in the initial stages of the pandemic, when the effects of mitigation steps were minimal, and employ a statistical framework that addresses the major sources of bias in estimating the generation interval distribution. We estimate a median generation interval of 7.9 days (95% CI 6.8-9) and an average of 9.7 days (95% CI 8.3-11.2), suggesting that the infectious period is much longer than previously thought. We demonstrate the effects of our updated estimate on the inference of effectiveness of case isolation and contact tracing.

## Methods

Data on serial intervals of transmission events were gathered from published and preprint literature, using a literature survey as described in the supplementary information. In order to control for biases introduced by later interventions, we focused on data from the early stages of the epidemic, when there were almost no cases identified outside China. Twelve relevant datasets were identified: ^10–12,16–24^. In total, the combined dataset contained 2,000 pairs, including duplicates. We cross-checked for duplicates in the combined dataset in three steps (see figure S1): First, we removed pairs containing the same “infector/infectee ID” (leaving 1685 pairs). Second, we looked at datasets containing sex and age information of the contacts and identified as duplicates those with matching sex, age, and symptom-onset date for both cases (identifying 931 unique transmission pairs in these sets). Lastly, we looked at the datasets not containing information regarding the sex and age of the cases (additional 406 pairs) and added to the dataset only pairs with symptom-onset dates that did not occur already in the in the first group (71 of the 406 cases were added, resulting in 1,002 transmission pairs in total). See figures S2 and S3 for a visualization of the datasets as a function of the symptom-onset date.

We estimated the unmitigated generation interval by focusing on the first period of transmission in China, thus minimizing the potential impacts of early interventions. To choose our analysis period, we relied on previous analyses of the early-outbreak and the timeline of interventions in Wuhan and mainland China. We quantified the forward serial-interval distributions based on the symptom-onset dates of the infector. We found that the mean forward serial interval stayed constant until around the 17th of January, 2020 and decreases gradually (Figure 2). The clear negative trend in the mean serial interval from January 17-18 onward matches the timing of the decrease in the effective reproduction number *R(t)* for domestic cases in Wuhan, China estimated by Lipsitch et al. ^*25*^. Large uncertainties in early serial interval data (before January 17) limited our ability to detect changes in the mean forward serial interval. Nonetheless, previous studies ^14,25,26^ found no clear signs of change in the growth of the epidemic prior to the period between the 16th and the 19th of January. Hence we assumed that the measures taken before January 20 such as changes in the China CDC emergency response levels ^18^ had a minimal effect on transmission. Notably, strict restrictions on mobility (lockdown) were imposed in Wuhan city on January 23, slowing down the spread of COVID-19 both inside and outside Wuhan ^26^. Hence, we would expect to see substantial reduction of the epidemic growth rate as well as shortened serial intervals starting with infectors infected a few days prior to this date. Based on this analysis, we used the transmission pairs for which the infector developed symptoms between December 12, 2019 - January 17, 2020 as our main dataset for estimating the unmitigated generation interval distribution. This dataset includes a total of 77 transmission pairs with a mean serial interval of 9.1 days (7.9-10.2 95% CIs), and a standard deviation of 5.2 days. This is substantially longer than the mean of 7.8 days suggested by Ali et al. ^10^ for the early period of the epidemic, at least in part because their estimates included infectors who developed symptoms up to January 22nd and were likely already subject to effects on mitigation strategies. Other studies that did not differentiate different stages of the epidemic estimated a much lower mean serial interval (4-6 days)^7,11,16,24^.

**Figure 2:**
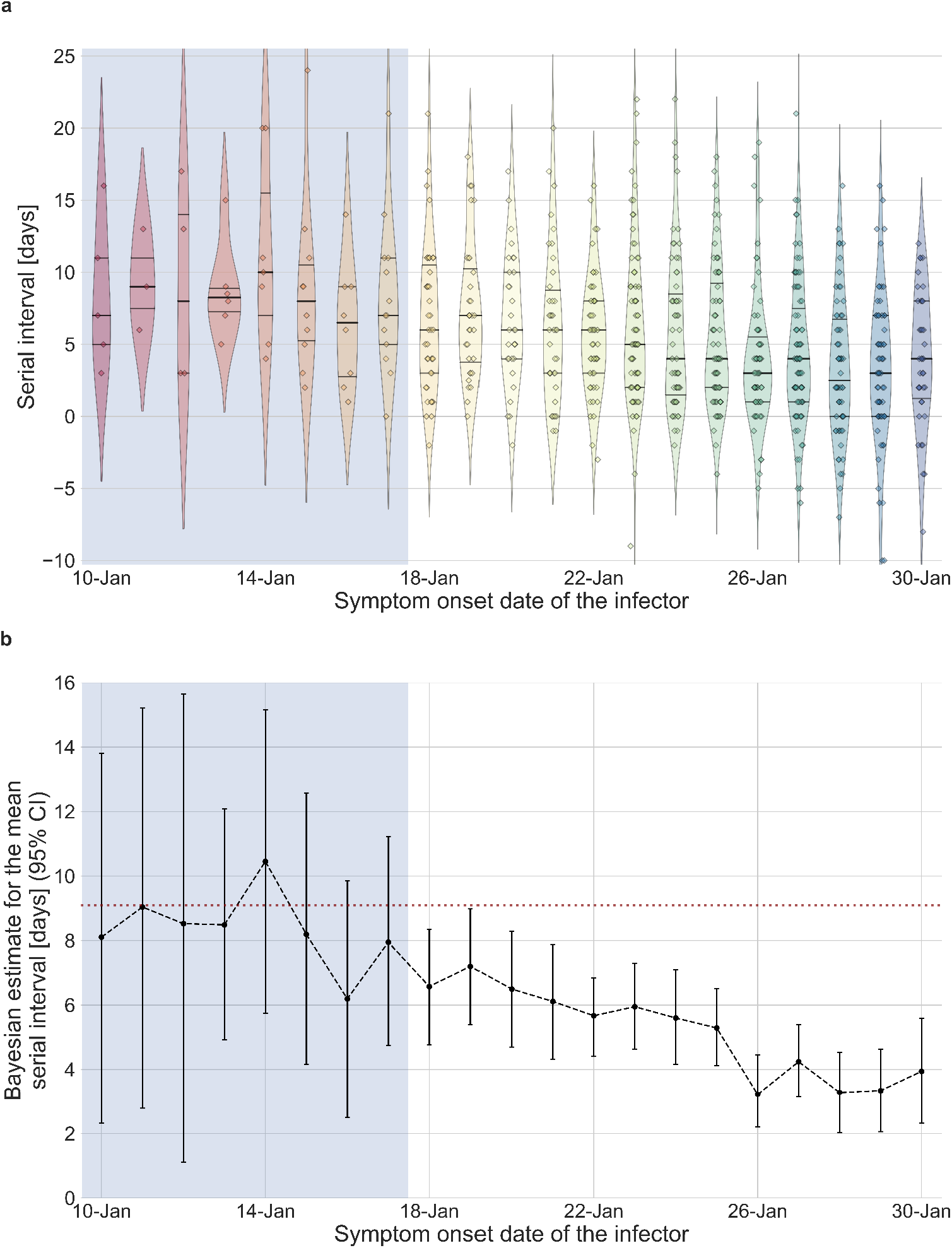
The serial interval dataset and the estimates of its mean during the early-outbreak period. **a**, The empirical distributions of forward serial intervals in the combined dataset, grouped based on the symptom-onset dates of the infectors and visualized using a violinplot. For pairs with uncertainty regarding the exact dates of symptom onset, we used a date in the middle of the uncertainty range. The violin shapes represent a kernel density estimation of the underlying distribution. The median and interquartile range (percentiles 25-75) are presented using dotted horizontal lines within the shape. The diamonds represent the data points for each of the dates of infector symptom onset. The dataset contains transmission pairs with infectors who developed symptoms from December 12 onward. Dates prior to January 10 are not shown as the data are too sparse b, The estimates of the mean serial interval, based on a parametric Bayesian inference (see Supplementary Information for details). The error bars represent the 95% confidence interval (CI) of the estimates. The dashed horizontal line represents the observed mean serial-interval for the period up to January 17. Dates up to January 17, 2020, are highlighted in both panels as they represent the period of unmitigated transmission.

Following Park et al. ^14,27^, our model incorporates the possible interaction of the generation-interval (τ_*g*_) with the incubation period of the infector(τ_*i*_) using a joint density function, denoted *h*(τ_*i*_, τ_*g*_).The use of a joint distribution allows us to consider a correlation between the two periods. For example, it is likely that infected individuals who develop symptoms later than average would also transmit later than average, given that viral load peaks around the time of symptom onset.

When the epidemic is in equilibrium (i.e., the incidence of infection remains constant over time) we can write down the probability density function *s*(τ|α_1_, α_2_) of observing an infector-infectee pair whose symptom-onset dates differ by a specific period (serial interval) τ. This probability density function is conditional on the infection time of the infector α_1_ and the infectee α_2_ relative to the symptom onset time of the infector. As described in Figure 1, if we define the symptom onset time of the infector as zero, this means that α_1_ < 0, and because the infector has to be infected before the infectee, this requires that α_1_ < α_2_. Assuming equilibrium conditions, *s*(τ_*i*_|α_1_, α_2_) is equal to the joint distribution describing the generation interval and the incubation period of the infector *h*(τ_*i*_, τ_*g*_), multiplied by the probability density function of the distribution of the infectee’s incubation period (denoted *l*(τ – α_2_).

This is a marginal distribution derived from *h* by integration over 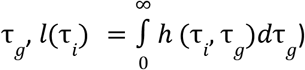 :

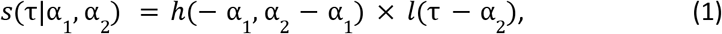

where τ is the serial interval, and α_1_, α_2_ are the infection times. The notations are further presented together with the definitions in Figure 1. As is shown in equation (1), the two distributions (*h*(τ_*i*_, τ_*g*_), *l*(τ_*i*2_)) depend on the relative infection times of both the infector and the infectee (α_1_ and α_2_). Although the exact time of infection is typically unknown, a possible exposure time window is provided in many cases. To compensate for the lack of information, the model integrates over all possible combinations of infector and infectee exposure times when estimating the parameters of the distribution from the observed serial intervals of the transmission pairs:

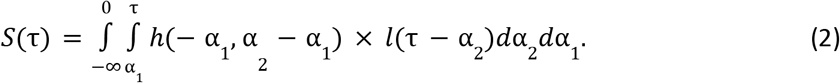

Most previous analyses of the serial-interval distributions of COVID-19 have relied on this model, which assumes a constant force of infection (i.e., the per capita rate at which susceptible individuals become infected). However, in the beginning of an epidemic, the number of infections (and therefore the force of infection) increases exponentially, creating a specific “backward” bias. When the force of infection is increasing exponentially, a cohort of infectors that developed symptoms at the same time is more likely to have been infected recently and thus to have shorter incubation periods, on average, than their infectees. Infectors with short incubation periods will also have short generation intervals due to their correlations, meaning that individuals who transmit early after infection are over-represented. It is important to correct for this bias by adding a factor 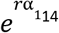:

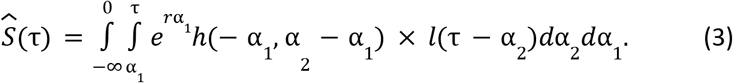

We used the incubation period distribution provided by a meta-analysis ^28 29^. The daily growth rates in the early outbreak period in Wuhan in particular and in the rest of China were estimated by another study ^30^ to be *r* = 0. 08 *d*^−1^ and *r* = 0. 10 *d*^−1^, respectively. In our main analysis, we used the growth rate measured for mainland China (*r* = 0. 10 *d*^−1^), taken as a mean growth rate representing the dynamic of the early outbreak relevant for most of the transmission pairs. We further present a sensitivity analysis for this parameter (see Results section). We note that daily growth rate estimates of 0.08-0.10 d^-1^ are lower than previous estimates in the range of 0.17-0.3 d^-1 31,32^ due to case ascertainment corrections ^31^. For the functional form of *h*, we used a bivariate log-normal distribution. Parameters for the incubation period were taken from the meta-analysis by Xin et al.^28^ leaving three free parameters: the shape and the scale of the log-normal distribution defining the generation-interval univariate distribution, and a correlation parameter (defined as the correlation between the logged incubation period and the logged generation interval). In order to test the sensitivity of our results to the choice of a log-normal distribution, we also considered the alternative form used in Ferretti et al. (2020)^3^ in supplementary Figure S9.

We then chose the parameters 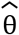 that maximize the likelihood of the observed serial intervals 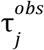 (the maximum likelihood estimate):

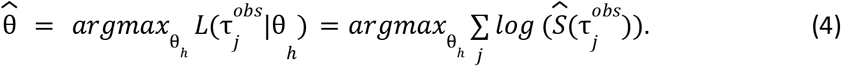

Sequential Least Squares Programming method, implemented in Python, was used to maximize the log-likelihood^33^. We calculated the uncertainties of the estimates using bootstrapping: the dataset was resampled with replacement (100 times for the main analysis and 100 times for sensitivity analyses) and processed via the maximum likelihood framework. In addition, the growth rate (*r*) was sampled from the uncertainty distribution found in a previous study of the early outbreak in China ^30^. We calculated the confidence interval based on the 95% quantiles of the bootstrapping results.

We conducted three primary sensitivity analyses to investigate potential biases in our approach. First, we tested how our estimate of the unmitigated generation-interval distribution is sensitive to our cutoff date assumption by varying it between January 11 and January 25. We note that using serial interval data from later dates are generally less reliable as they are affected by mitigation measures, which prevent late transmissions. Second, we considered the possibility that long serial intervals may be caused by omission of intermediate infections in multiple chains of transmission, which in turn would lead to overestimation of the mean serial and generation intervals. Thus, we tried to refit our model after removing long serial intervals from the data (by varying the maximum serial interval between 14 and 24 days). Finally, we considered the possibility that the lack of negative serial intervals in early serial interval data might have been caused by the incorrect determination of the direction of transmission, especially given limited information about presymptomatic transmission in the beginning of the pandemic. To test for potential biases, we refitted our model after switching the direction of transmission among randomly selected infector-infectee pairs by varying the number of pairs switched (2, 4, 6, or 8 pairs out of 77) and the maximal serial interval for which order switching is allowed (3, 5, or 7 days). For each combination, the analysis was run 30 times with randomly sampled infector-infectee pairs.

Furthermore, we tested other possible sensitivities of the data to biases based on location of infection, or the literature source of the data. To test the sensitivity to infection location, we stratified the dataset by where the infectors were infected (Wuhan vs outside of Wuhan) as detailed in the supplementary information. To test for sensitivity to any specific literature source, we repeated the analysis while removing one dataset at a time, including all the transmission events that were duplicated also in other datasets (defined by the infector and infectee ID).

Additional sensitivity analyses were performed for other aspects of the analysis. The effect of the assumed growth rate was assessed by varying it between 0.04-0.16 d^-1^. Furthermore, the sensitivity of the results to the choice of the lognormal bivariate distribution model was tested by comparison with another model distribution given in Ferretti et al. ^3^ (see supplementary material for full details).

## Results

We inferred the unmitigated generation-interval distribution of SARS-CoV-2 transmission based on an integrative curated dataset, which focuses on the early-outbreak period in China.

We used the maximum likelihood framework to estimate the parameters of the joint bivariate distribution of the generation interval and the incubation period, assuming a known incubation-period distribution ^28^ with a mean of 6.3 days and a standard deviation of 3.6 days. We estimate that the unmitigated generation-interval distribution has a median of 7.9 days (95% confidence interval (CI): 6.8-9), a mean of 9.7 (95% CI: 8.3-11.2) days and standard deviation of 6.9 (95% CI: 4.3-10.1) days. Furthermore, we estimate a correlation parameter (see Methods) of 0.75 (95% CI: 0.5-0.9). Our estimates are robust to the choice of data sources used in the analysis included (Figure S5).

We note that the estimated mean generation-interval is longer than the observed mean serial-interval (9.1 days) of the period in question--in contrast to the common assumption that the mean generation and serial intervals are identical, the mean forward serial interval, which we observe, can systematically differ from the mean generation interval, which we aim to estimate here, due to dynamical effects. During the exponential growth phase, the mean incubation period of the infectors is expected to be shorter than the mean incubation period of the infectee - this effect causes the mean forward serial interval to become longer than the mean forward generation interval of the cohorts that developed symptoms during the study period. However, these cohorts of individuals with short incubation periods will also have short forward generation (and therefore serial) intervals due to their correlations. The intrinsic generation intervals we estimate are longer because they are not conditioned on short incubation periods ^14^.

The joint bivariate distribution and its marginal distributions are shown in Figure 3a. We find that our framework is able to properly reproduce the realized serial interval distribution given the growth rate in the early stages of the outbreak in Wuhan, China (Figure 3b). Using the inferred bivariate distribution, we derived the distribution of time from onset of symptoms to transmission (TOST), shown in Figure S4. The negative side of this distribution gives the pre-symptomatic transmission, which constitutes ≈20% (95% CI: 6%-32%) of total transmission.

**Figure 3:**
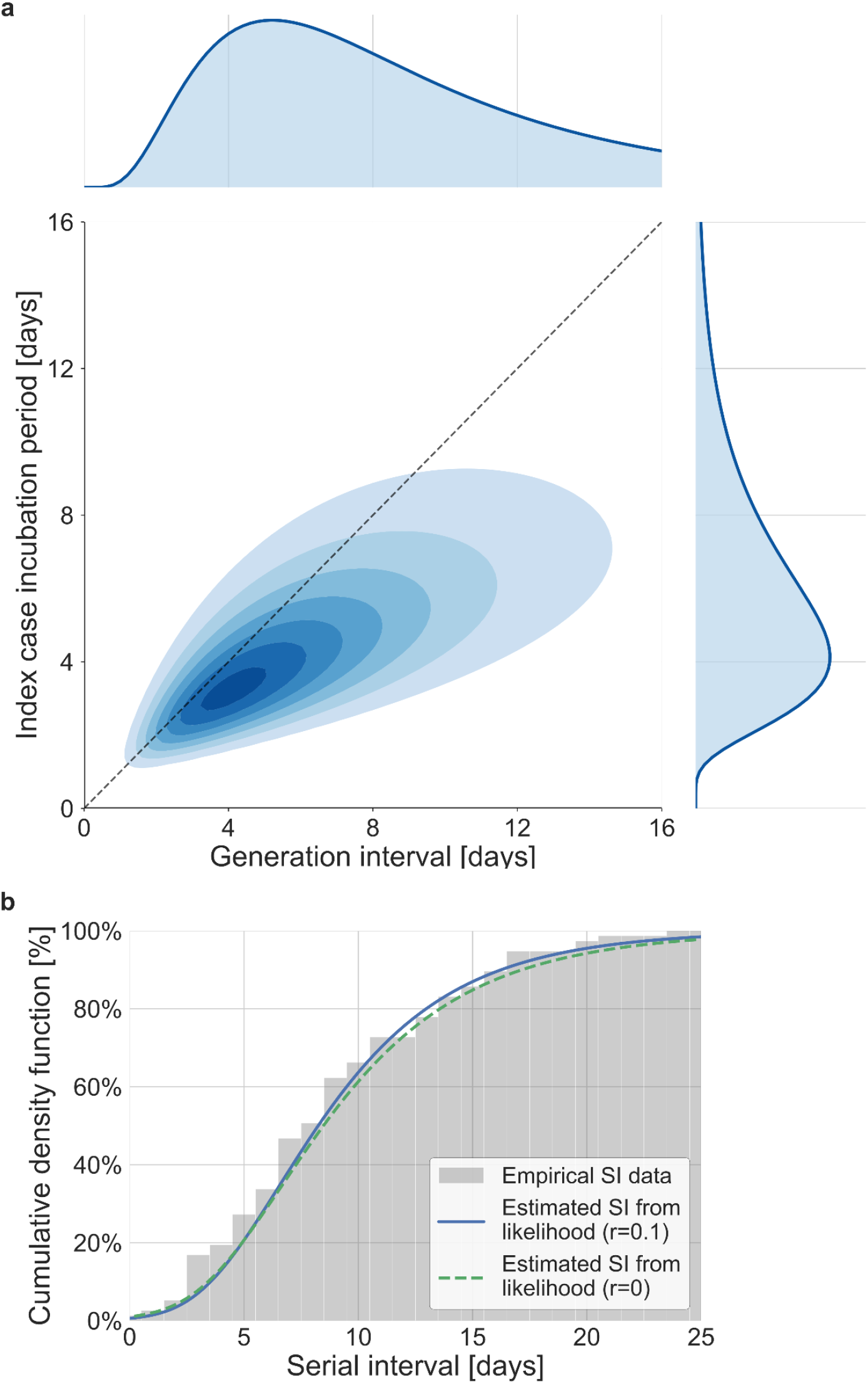
The joint distribution of generation interval and incubation period. Representations of the inferred joint distribution results are based on maximum likelihood analysis. **a**, The joint bivariate distribution (bottom left graph), shown as contours over the plane of generation intervals (x-axis) and incubation period distribution (y-axis). The correlation parameter (in log space, see Methods) was found to be 0.75(0.5-0.9 95% CI). The panel also shows the univariate components of the joint distribution: the generation interval distribution (top graph, sharing the same x-axis) and the incubation period distribution (bottom right graph, sharing the same y-axis). The incubation period distribution was assumed to follow a log-normal distribution with a shape parameter of 0.53 and a scale parameter of 5.5 days, following Xin et al. ^28^. The dashed grey diagonal line describes equal incubation period and generation interval (TOST equal to zero). Left of this line could be found the pre-symptomatic fraction of transmission. **b**, Cumulative histogram of the empirical serial intervals and the parametric distribution derived from the maximum likelihood joint distribution. The estimated serial interval distribution was derived using the likelihood calculation given the reported growth rate of r=0.1/d ^30^. For comparison the dash line represents the intrinsic serial interval distribution, estimated by equation (2) with the parameters derived from the maximum likelihood analysis.

A comparison with the current available estimates of the generation interval distribution ^3,7,11^ reveals that the inferred distribution has a heavier (right) tail (Figure 4) and a higher median (7.9 days compared to 5.4-5.8 days) and standard deviation (6.9 days compared to 3.3-3.9 days). For example, the gamma distribution assumed by Johansson et al. (2021) to give an infectious period of about 10 days (and a peak at 5 days) for the analysis of quarantine and isolation policies has a far smaller tail. One way to quantify the difference in the tails of the different estimates is by comparing the proportion of transmission after a certain time-point. When comparing the proportion of transmission after day 14, there are clear differences from previously reported distributions. The distributions of Ferretti et al., He et al. and Sun et al. indicate a residual fraction of transmission after 14 days of 2%-4%, while the distribution assumed by Johansson et al. indicates only 0.2%. In contrast, our inferred generation-interval distribution predicts that about 18.5% (95% CI of 10%-25%) of the transmission occurs after 14 days in the unmitigated scenario.

**Figure 4:**
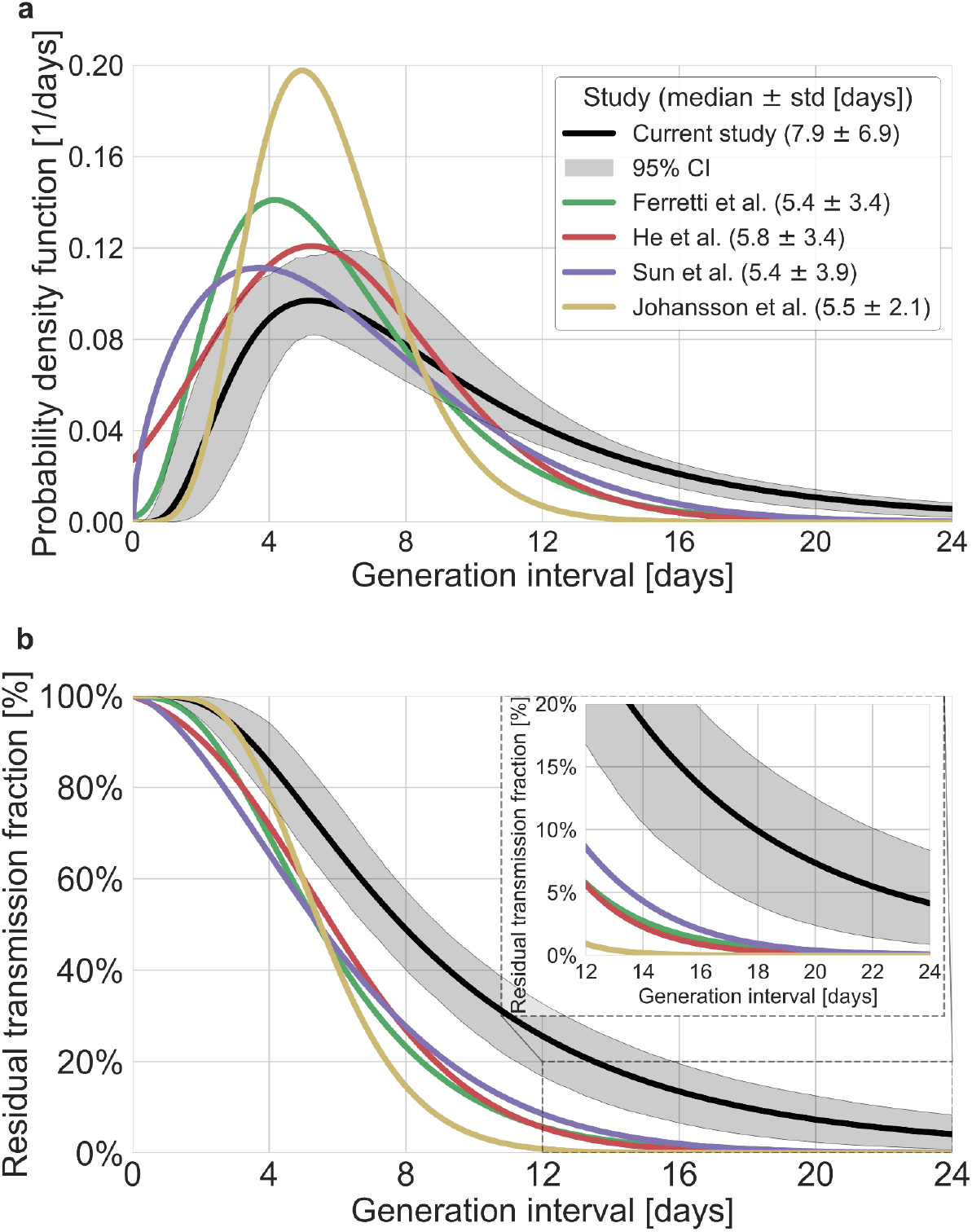
Comparison of the mean generation interval distribution with those of previous studies. The generation-interval distribution inferred by maximum likelihood presented alongside available estimates from the literature ^3,7,9,11,34^. **a**, The probability density functions of the distributions. The legend reports the median and standard deviation of each of the distributions. **b**, The survival function of the generation interval distribution, defined as the complement of the cumulative distribution, representing the residual fraction of transmission after a designated time since infection. The inset shows a zoom-in on the period of 10-24 days after exposure, a period in which there is a substantial difference between the current estimate and those from previous studies. The highlighted area represents the 95% confidence interval of the maximum likelihood estimate.

In addition to the possible dynamical and statistical biases considered in our analysis, the resulting wide generation interval distribution might be affected by biases in the data collection process as detailed in the Introduction and Method sections. The estimated generation-interval distributions were sensitive to the cut-off date with an estimated median of 6.5-8 days and estimated means of 7-10 days for periods ending on January 16 to January 19, 2020 (Figure 5a-c and Figure S6).

**Figure 5:**
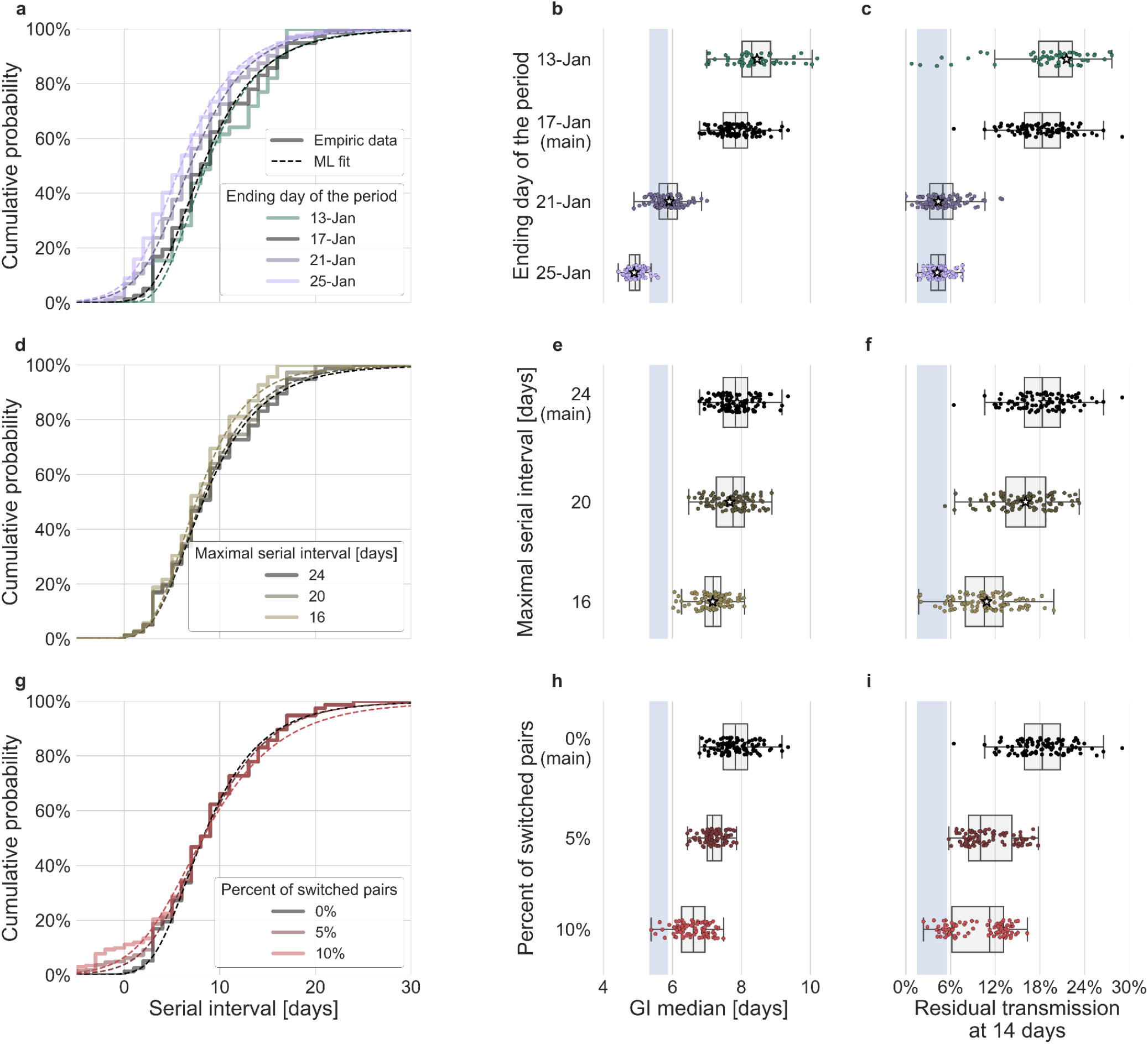
Sensitivity analyses of the inferred generation interval. A comparison of the results of sensitivity analysis to three factors: the period chosen to represent the unmitigated transmission (**a-c**), the inclusion of the longest serial intervals in the dataset (**d-f**), and the ordering of the transmission pairs(**g-i**). **a, d, g**, Cumulative histogram of the empirical serial intervals and the parametric distribution derived from the maximum likelihood joint distribution. The estimated serial interval distribution was derived using the likelihood calculation given the reported growth rate of r=0.1/d ^30^, **e, h**, Best estimates and distributions of the resulting median of the inferred generation interval distribution. Best estimates are marked by a black star. Ranges are given as boxplots. The box represents the interquartile range (percentiles 25-75) and the whiskers represent the maximal range of the distribution apart from outliers (defined as data points exceeding the interquartile range by a factor of 1.5). Each dot represents a single bootstrapping iteration. The blue shaded region represents the values from previous studies ^3,7,11 30^, **f, i**, Best estimates and distributions of the resulting residual transmission at 14 days since infection derived from the inferred generation interval distribution. The best estimates and ranges are shown in the same manner as the distribution parameters in panels^30^, **e, h**.

Switching the order of some of the transmission pairs caused a decrease in both the median and mean of the generation interval, as well as a decrease in the correlation parameter (Figure 5g-i, Figure S13). The sensitivity analysis to high serial-interval values caused a slight decrease in the mean generation interval, but still resulted in a wide distribution. Removing the transmission pairs with the highest serial intervals from the dataset caused a small decrease in the generation interval distribution. For a removal of the top 10% values, the inferred distribution has a median of 7.2 days and a mean of 8.3 days (Figure 5d-f, Figure S14). As switching the direction of transmission among randomly selected infector-infectee pairs gives negative serial intervals (and thus lower mean serial interval) a decrease in the mean generation interval distribution was expected. However, even when reordering 10% of the pairs the distribution is wide: for example, the median of bootstrap estimates for the median generation interval is 7.2 days (Figure 5h). These bootstrap estimates also yield substantial residual transmission at 14 days (Figure 5i).

Another factor of uncertainty in the estimate is the growth rate we assume for the inference of the distribution. Changing the assumed growth rate during this period had very little effect on the results, with estimated mean increasing from 9.5-9.7 days, as assumed growth rates decreased from 0.16-0.04 d^-1^. These sensitivity analyses demonstrate the robustness of our conclusion: the unmitigated generation-interval distribution is likely wider than previously thought.

Finally, to quantify the effect of our estimated generation interval distribution on the estimates of the basic reproduction number R_0_, we use the growth rate estimated in a recent study of the early outbreak dynamics in China ^30^. Combining our estimated generation interval distribution with the early growth rate ^5^ we find R_0_ to be 2.2 with a confidence interval of 1.9-2.7 (Figure S7).

## Discussion

In this work, we assembled transmission-pair data from 12 datasets representing the early-outbreak period in China, and modeled the relationship between disease transmission and symptom onset using a bivariate log-normal distribution. By applying a maximum-likelihood framework, we found that the unmitigated generation-interval distribution has a heavier right tail than previously estimated ^3,7,11^, corresponding to a larger mean and standard deviation. The bias in the previous estimates likely reflects the effects of mitigation steps, such as quarantine of exposed individuals, as well as changes in awareness-driven behavior, such as faster self-isolation after symptom onset, that prevent transmission during late stages of infection. These sources of biases were not fully accounted for in previous estimates, leading to substantial underestimation of the generation-interval distribution.

Furthermore, accounting for potential correlations between the incubation period and the generation interval provided a better estimate of the proportion of pre-symptomatic transmission. Our results suggest that, on average, only ≈20% (6%-32%) of the unmitigated transmission happens before symptoms appear, lower than commonly stated values that already include mitigation effects (40-60% ^3,7^). When mitigation strategies are introduced, we would expect the amount of post-symptomatic transmission to decrease, leading to an increase in the fraction of pre-symptomatic transmission. Thus, it is not surprising that our estimate of the proportion of pre-symptomatic transmission is lower than previous estimates that looked at a later period ^3,7^. Furthemore, our results match the trend shown by Sun et al. ^7^, in which the faster isolation of cases increases the pre-symptomatic fraction of transmission and shortens the mean generation interval.

To check whether these results are sensitive to our choice of using a bivariate lognormal distribution to characterize the joint distribution of the generation interval and the incubation period, we repeated our analysis using a different functional form using an adjusted logistic TOST model following ^3^ (see supplementary for details). Both models estimate large means and standard deviations of the generation intervals, and a low proportion of pre-symptomatic transmission for the current dataset. Applying both models to the data from Ferretti et al. ^3^ produced similar distributions with lower estimates for the mean generation interval and higher per-symptomatic proportion (Figure S9). This indicates that the results presented in this study are a product of the focus on the data prior to mitigation steps, in combination with the correction for the growth of the epidemic.

Our analysis relies on datasets of transmission pairs gathered from previously published studies and thus has several limitations that are difficult to correct for. Transmission pairs data can be prone to incorrect identification of transmission pairs, including the direction of transmission. In particular, presymptomatic transmission can cause infectors to develop symptoms after their infectees, making it difficult to identify who infected whom. Data from the early outbreak might also be sensitive to ascertainment and reporting biases. For example, people who transmit asymptomatically might not be identified. Moreover, when multiple potential infectors are present, an individual who developed symptoms close to when the infectee became infected is more likely to be identified as the infector. These biases might increase the estimated correlation of the incubation period and the period of infectiousness. We have tried to deal with these biases by using a bootstrapping approach, in which some data points are omitted in each bootstrap sample. The relatively narrow ranges of uncertainty suggest that the results are not very sensitive to specific transmission pairs data points being included in the analysis. We also performed a thorough sensitivity analysis to address several of the potential biases such as the determination of period corresponding to unmitigated transmission, the inclusion of long serial intervals in the dataset, and the incorrect orderings of transmission pairs (see Method). The sensitivity analysis shows that although these potential biases can decrease the inferred generation interval distribution, our main conclusions about the long unmitigated generation intervals remained robust with a high median and residual transmission after 14 days compared to previous estimate (Figure 5).

Our estimates of the unmitigated generation-interval distribution can inform quarantine policy. The tail of the survival function (Figure 4b) indicates that individuals released from quarantine at day 14 still have, on average, ≈18% of their transmission potential. We also found a strong correlation of the incubation period with the generation interval, accentuating the importance of quickly isolating individuals as soon as they show symptoms.

Determining the optimal period of quarantine for individuals exposed to COVID-19 is hard, as it needs to balance the prevention of further transmission with personal and economic costs of longer quarantine. It is important to consider the basic risk of transmission underlying those considerations, by looking at the distribution of infectiousness in the absence of mitigation measures. Johansson et al.’s (2021) estimates for the residual transmission across different quarantine policies (e.g., with and without testing before release) have served as the basis for recent recommendations by the U.S. Centers for Disease Control and Prevention^35^ for a 10-day quarantine period (without PCR testing) for exposed individuals. As can be seen in Figure 4b, our results suggest that this analysis underestimates the residual transmission after 10 days by an order of magnitude for the average individual (35% of the transmission versus 4%). One of the first and ongoing policies from mitigating transmission is mandatory self-isolation for individuals developing COVID-19 related symptoms ^9,36^. We estimate a strong correlation of incubation period and infectiousness, enhancing the contribution of self-isolation to transmission prevention. However, even when considering self-isolation of 70% of individuals immediately upon symptoms, as Johansson et al. ^9^ assumed in their analysis, we still find a residual transmission of 11.8% compared to 1.3% in Johansson et al.’s estimates (Figure S8). It should be noted that using PCR or rapid tests during quarantine has a dramatic potential to reduce the residual transmission after quarantine, and hence is required in many countries. The current study does not analyse the possible benefits of such policies, but only of self-isolation by individuals who developed COVID-19 symptoms.

The basic reproduction number R_0_ estimates derived here are close to reported values from early in the epidemic value^18,37–39^.

The current analysis provides an updated benchmark for the unmitigated profile of SARS-CoV-2 infectiousness. Furthermore, with the emergence of new variants of concern, which may exhibit altered transmission dynamics than previously dominant wild type strains ^40^, future studies could use our framework to update estimates of the generation interval for these emerging strains even under mitigation conditions and with inference of the correlation to the incubation period.

Taken together, our results demonstrate the importance of considering possible biases in the serial-interval data used for estimating the generation-interval distribution, as well as the underlying assumptions made when estimating the distribution from the source data. Our analysis provides a view of the infectiousness profile of an infected individual in absence of mitigation steps, which is a key ingredient of many models used for guiding policy.

## Data Availability

All data produced are available online at https://gitlab.com/milo-lab-public/the-unmitigated-profile-of-covid-19-infectiousness

## Acknowledgments

We would like to thank David Champredon and David Earn for valuable feedback on this manuscript. Funding: Ben B. and Joyce E. Eisenberg Foundation, The Weizmann CoronaVirus Fund (R.M.), the Israeli Council for Higher Education (CHE) via the Weizmann Data Science Research Center, and by a research grant from the Estate of Tully and Michele Plesser (R.S.). R.M. is the Charles and Louise Gartner Professional Chair. Jonathan Dushoff is supported by the Canadian Institutes of Health Research. Y.M.B. is an Azrieli Fellow

## Data and code availability

All study data are included in the article, SI appendix, and Dataset S1.

All code is available in Jupyter notebooks found in https://gitlab.com/milo-lab-public/the-unmitigated-profile-of-covid-19-infectiousness

## Supplemental Figures

**Figure S1:**
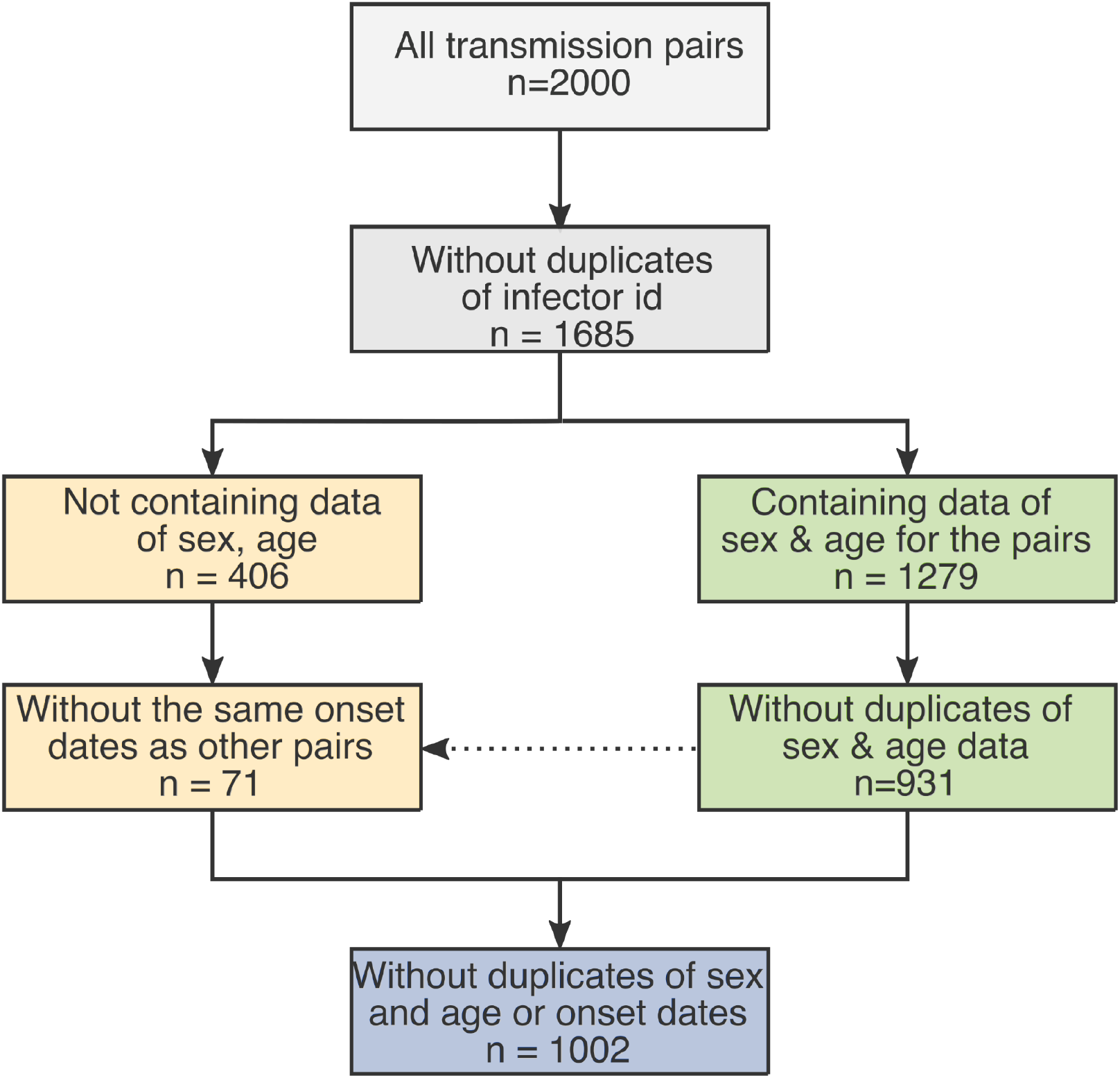
The obtained dataset of transmission pairs. The merged datasets were filtered to remove duplicates in three stages: first removal of transmission pairs with the same infector and infectee id. Second, identification of duplicates sharring the same symptom-onset dates as well as sex and age information. Lastly, transmission pairs without sex and age information were added only if their symptom-onset dates didn’t already occur in the dataset.

**Figure S2:**
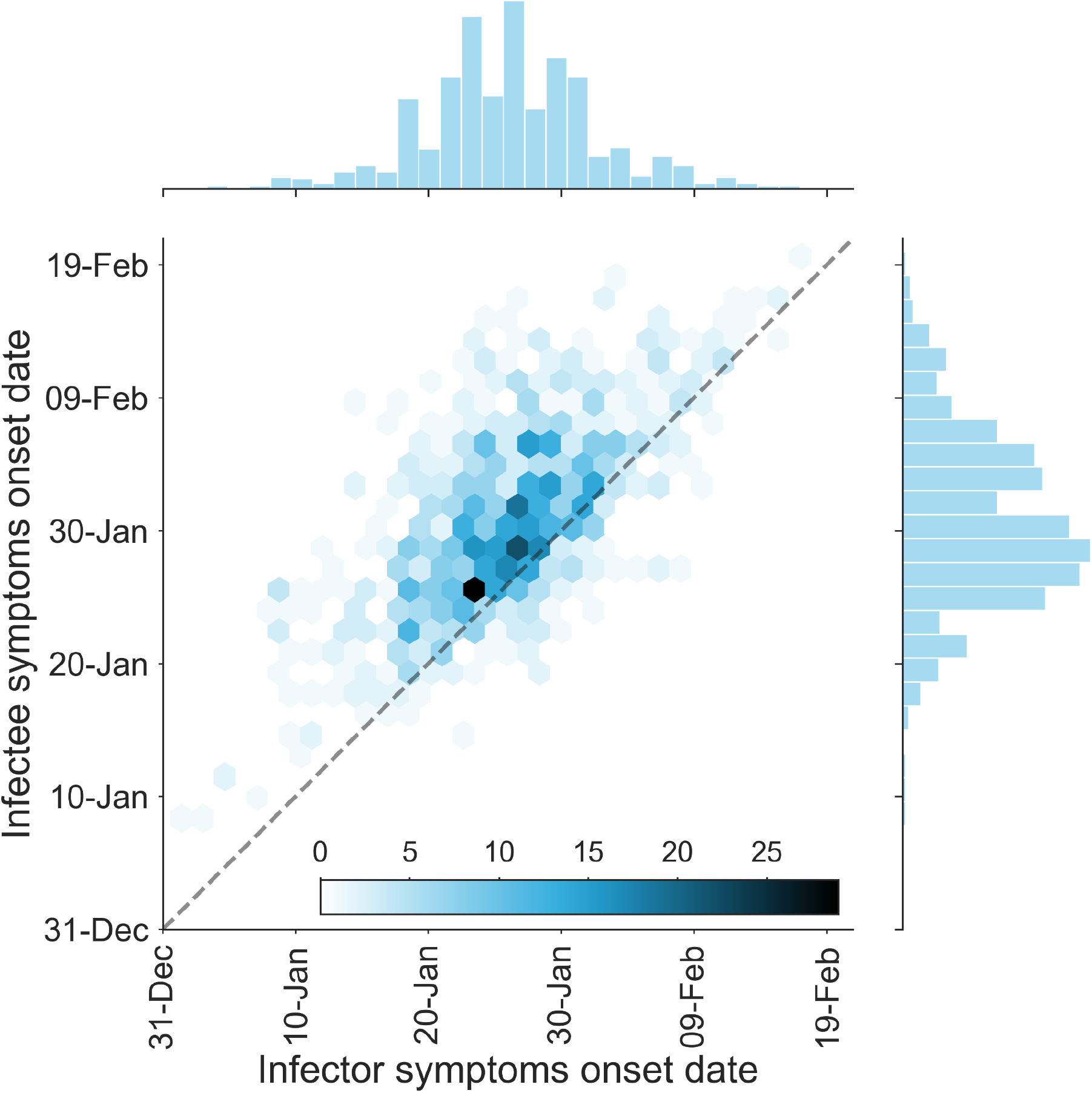
The dataset of transmission pairs - infectee symptoms onset date vs. infector symptoms onset date. The observed bivariate distribution of time of symptoms development is shown via a hexbin graph. The plane is divided into hexagons that are colored according to the number of data points in the dataset they represent. The marginal empirical distributions are shown using a histogram on the sides. The dotted line represents data points for which the symptoms’ onset date of the infector is the same as that of the infectee.

**Figure S3:**
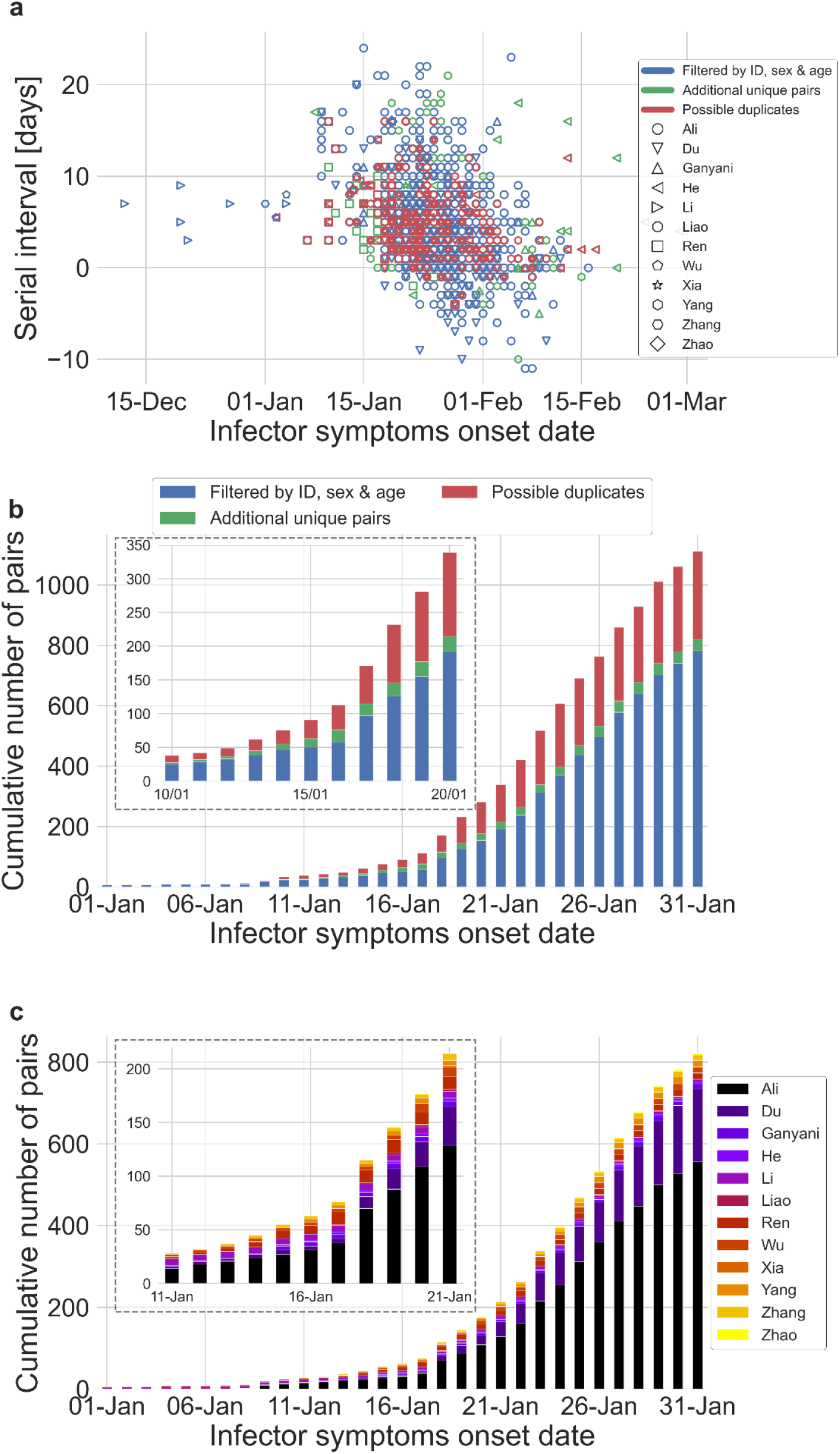
The dataset of transmission pairs as a function of the infector symptoms onset date. **a**, Scatter plot of serial intervals plotted against the symptoms onset date of the infector. The three levels of filtering are color-coded, while the shape of the marker represents the reference from which the data was taken. **b**, The cumulative number of cases as a function of the infector symptoms onset date, where the data is divided between the three levels of filtration. The inset focuses on the period that at its end interventions were made. **c**, The cumulative number of cases as a function of the infector symptoms onset date, where the data is divided between the data sources. The dataset is shown after filtrating by ID, sex & age and the addition of unique pairs (no duplicates). The inset focuses on the period that at its end interventions were made.

**Figure S4:**
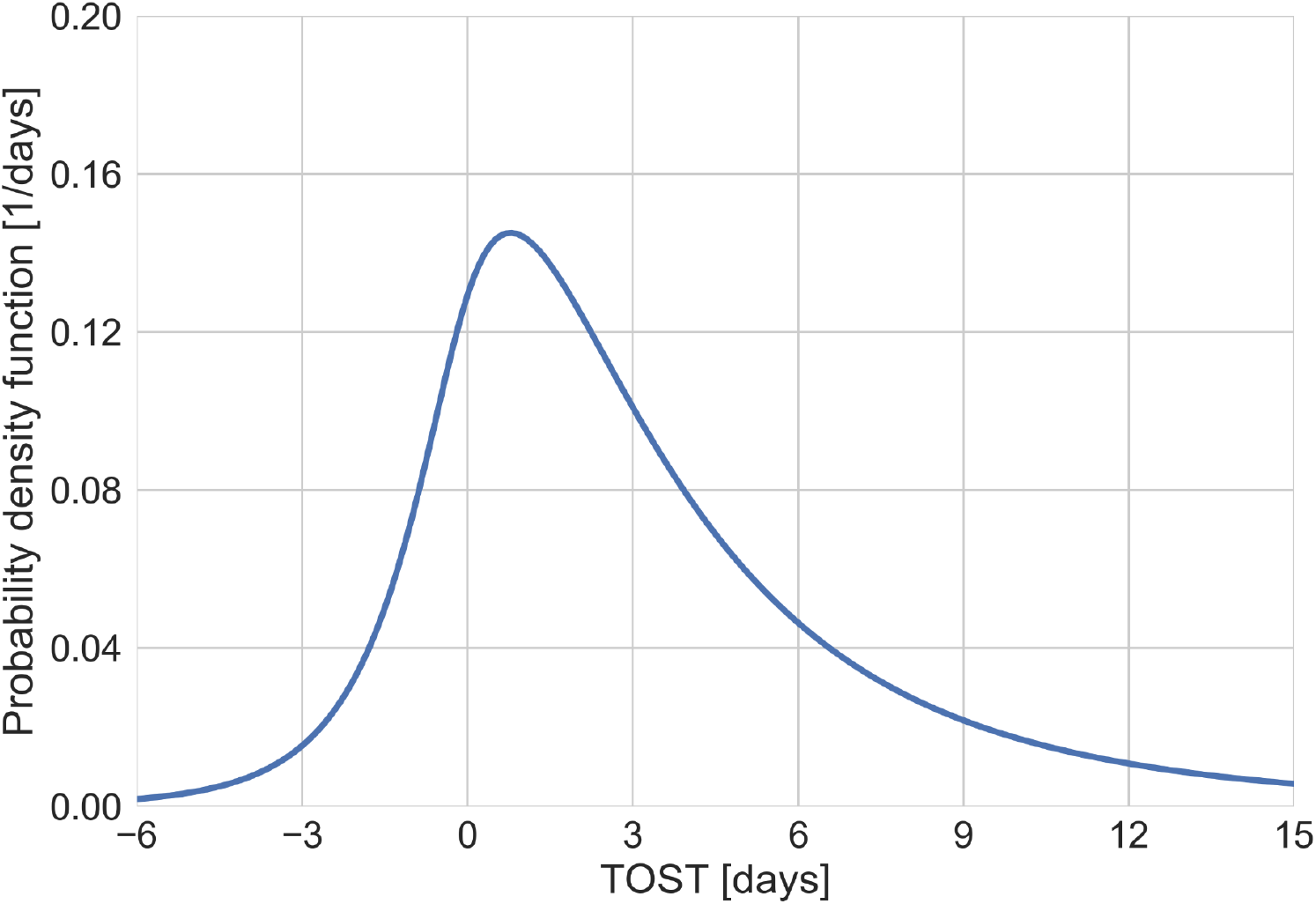
The distribution of time from onset of symptoms to transmission (TOST). Derived from the joint bivariate lognormal distribution.

**Figure S5:**
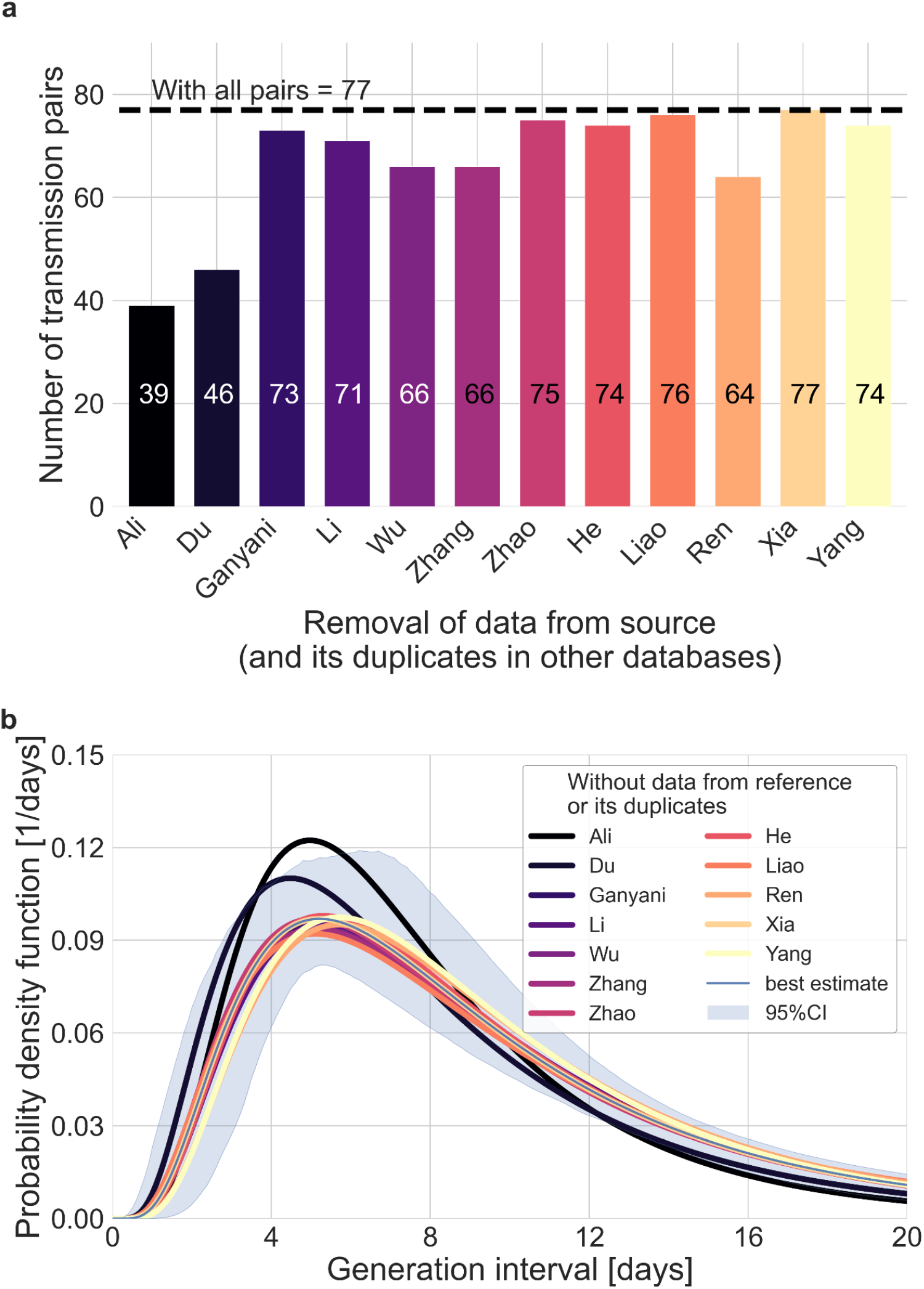
Sensitivity analysis regarding the inclusion of a dataset from a specific source. Beginning with the complete dataset of transmission pairs with infector onset until 17 January, 2020 (after filtering by ID, sex & age and adding unique pairs, see S2), partial datasets were created by omitting all transmission pairs from each source of data and all its duplicates in the other datasets. **a**, The number of transmission pairs for each of the partial datasets, excluding pairs from a specific source dataset and their duplicates in other datasets. **b**, Maximum likelihood estimates of the bivariate incubation period and generation interval distribution. The uncertainty range of the maximum likelihood estimate of the complete dataset is also shown for comparison.

**Figure S6:**
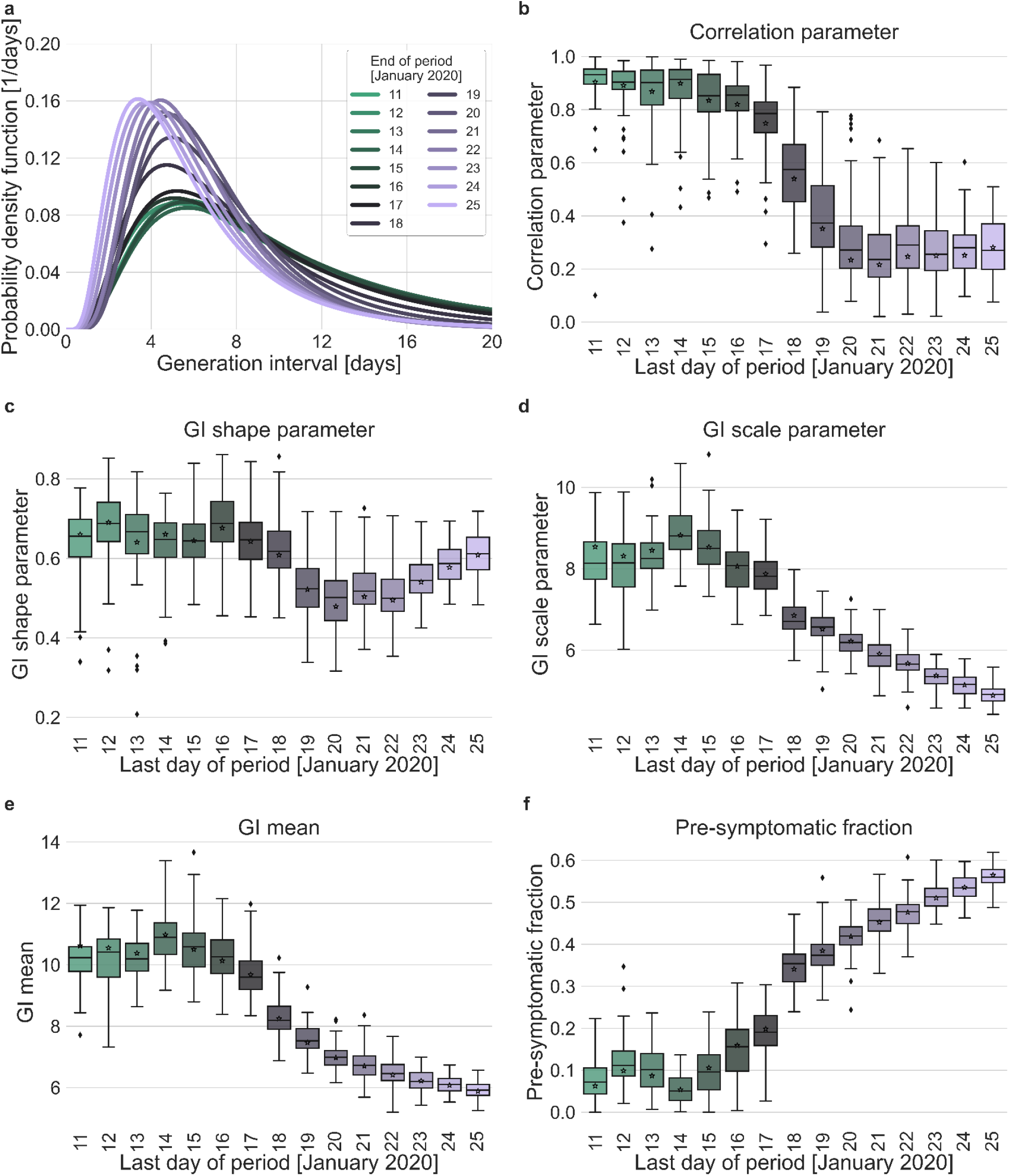
Sensitivity analysis regarding the choice of period for analysis. Maximum likelihood estimates of the bivariate incubation period and generation interval distribution were made for datasets containing the transmission pairs with infector onset date up to a specific date. Best estimates were derived for each of the datasets. Uncertainty estimates were derived by bootstrapping, through sampling with replacement from the dataset and sampling from the distribution of growth rates ^30^. **a**, Best estimate for the generation interval distribution probability density function for periods ending at dates in the range of 11-25 January, 2020. **b-d**, Best estimates and distributions of the resulting parameters of the bivariate distribution of incubation period and generation interval. Best estimates are marked by a black star. Ranges are given as boxplots. The box represents the interquartile range (percentiles 25-75) and the whiskers represent the maximal range of the distribution apart from outliers (defined as data points exceeding the interquartile range by a factor of 1.5). **e-f**,. The mean generation interval and the fraction of pre-symptomatic transmission, derived from the results. The best estimates and ranges are shown in the same manners as the distribution parameters in panels **b-d**.

**Figure S7:**
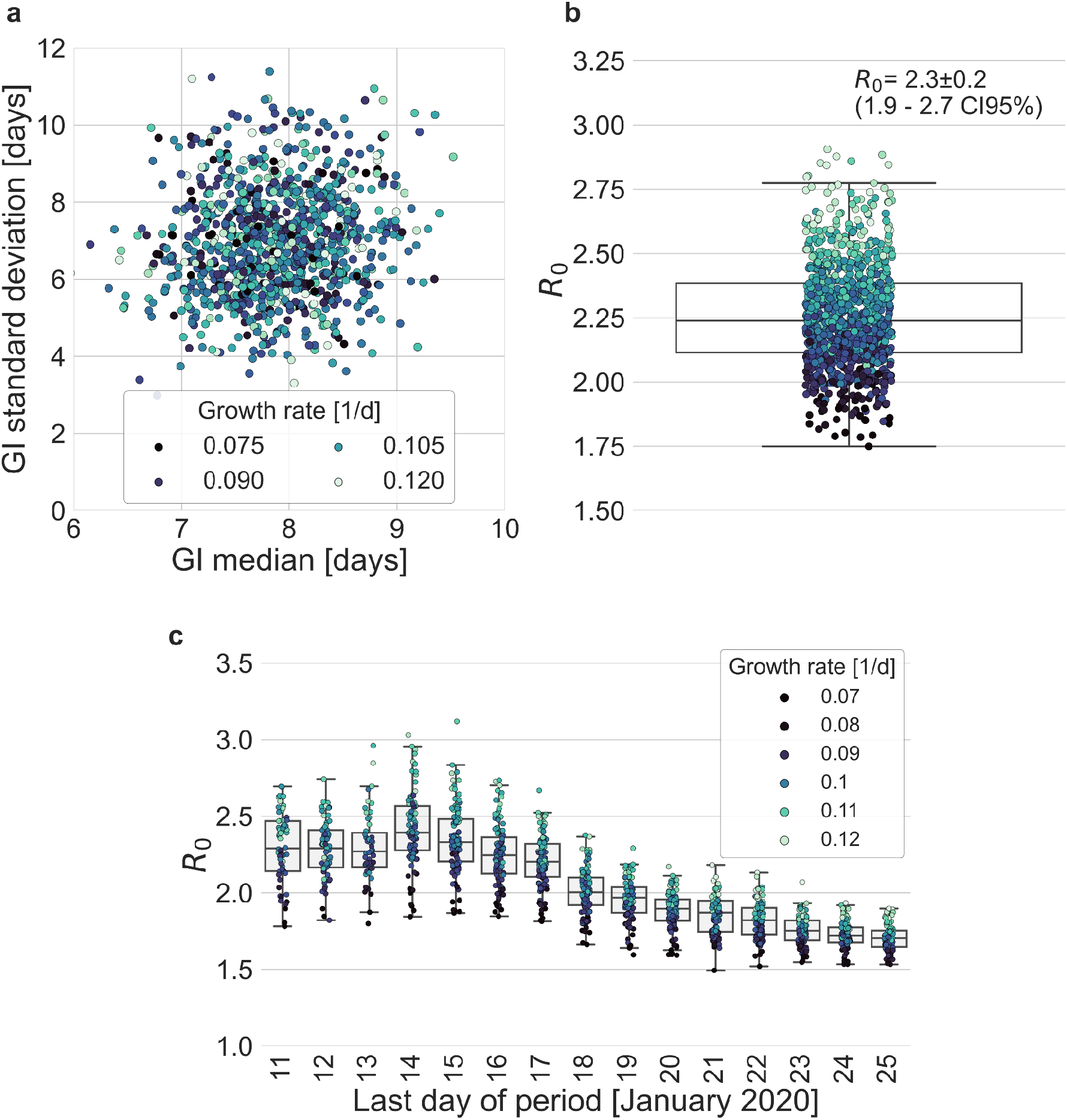
Estimates of R_0_ based on the inferred generation interval distribution. **a-b**, Bootstrapping results of the parameters of the generation interval distribution and the resulting estimates for R_0_. In the process of bootstrapping, the dataset of 77 transmission pairs was resampled with returns. In addition, the growth rate (r) was sampled from the distribution found in a recent study ^30^ **a**, Estimates of the mean and standard deviation of the generation interval. Each point represents the maximum likelihood estimate for a single run in a bootstrap process. The point was colored according to the sampled growth rate. **b**, The distribution of estimates of R_0_ derived from the generation interval distribution and growth rate. The box represents the interquartile range (percentiles 25-75) and the whiskers represent the maximal range of the distribution apart from outliers (defined as data points exceeding the interquartile range by a factor of 1.5). The mean (with its 95% confidence interval) and the standard deviation is given in the legend. The points are colored according to the sample growth rate, as in panel a. **c**, The dependence of R_0_ estimates on the period taken in the analysis. The boxes represent the interquartile range (percentiles 25-75) and the whiskers represent the maximal range of the distribution apart from outliers (defined as data points exceeding the interquartile range by a factor of 1.5). The points are colored according to the sample growth rate, as described in the legend.

**Figure S8:**
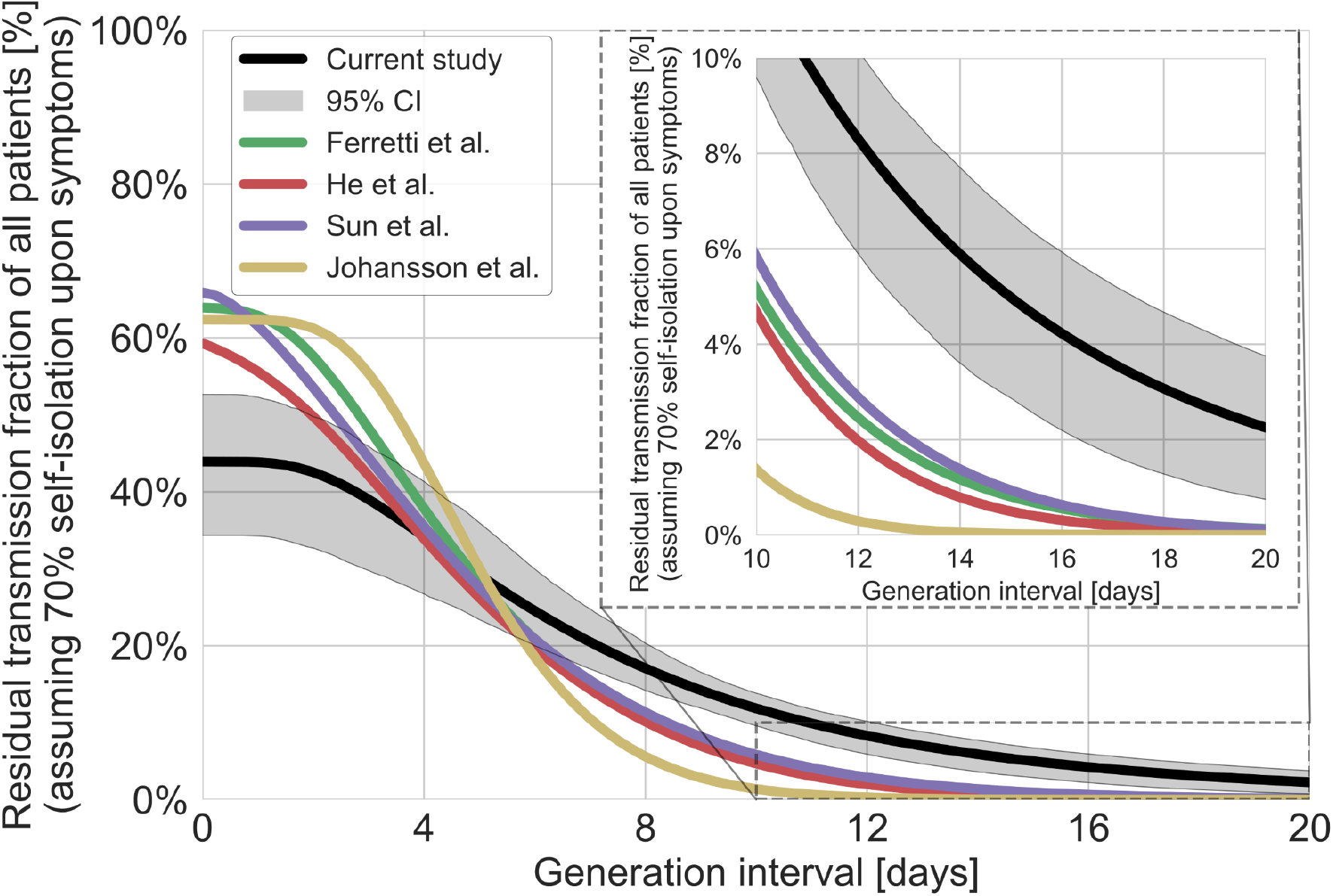
The residual transmission accounting for self-isolation. The residual transmission under the conjecture that 70% of individuals self-isolate upon the development of symptoms was calculated as a weighted average of the regular survival function (shown in Figure 4b) and the residual transmission conditioned on the self-isolation function. This analysis was performed for the distribution of generation intervals inferred by maximum likelihood as well as for best available estimates from the literature ^3,7,9,11,34^. The residual transmission conditioned on self-isolation function is calculated through the integration of the bivariate distribution of incubation period and generation interval, on the relevant quadrant (the probability summed on incubation and generation interval greater than a specific value). The inset shows a zoom-in on the period of 10-20 days after exposure, a period in which there is a substantial difference between the current estimate and those from previous studies. The highlighted area represents the 95% confidence of the maximum likelihood estimate.

**Figure S9:**
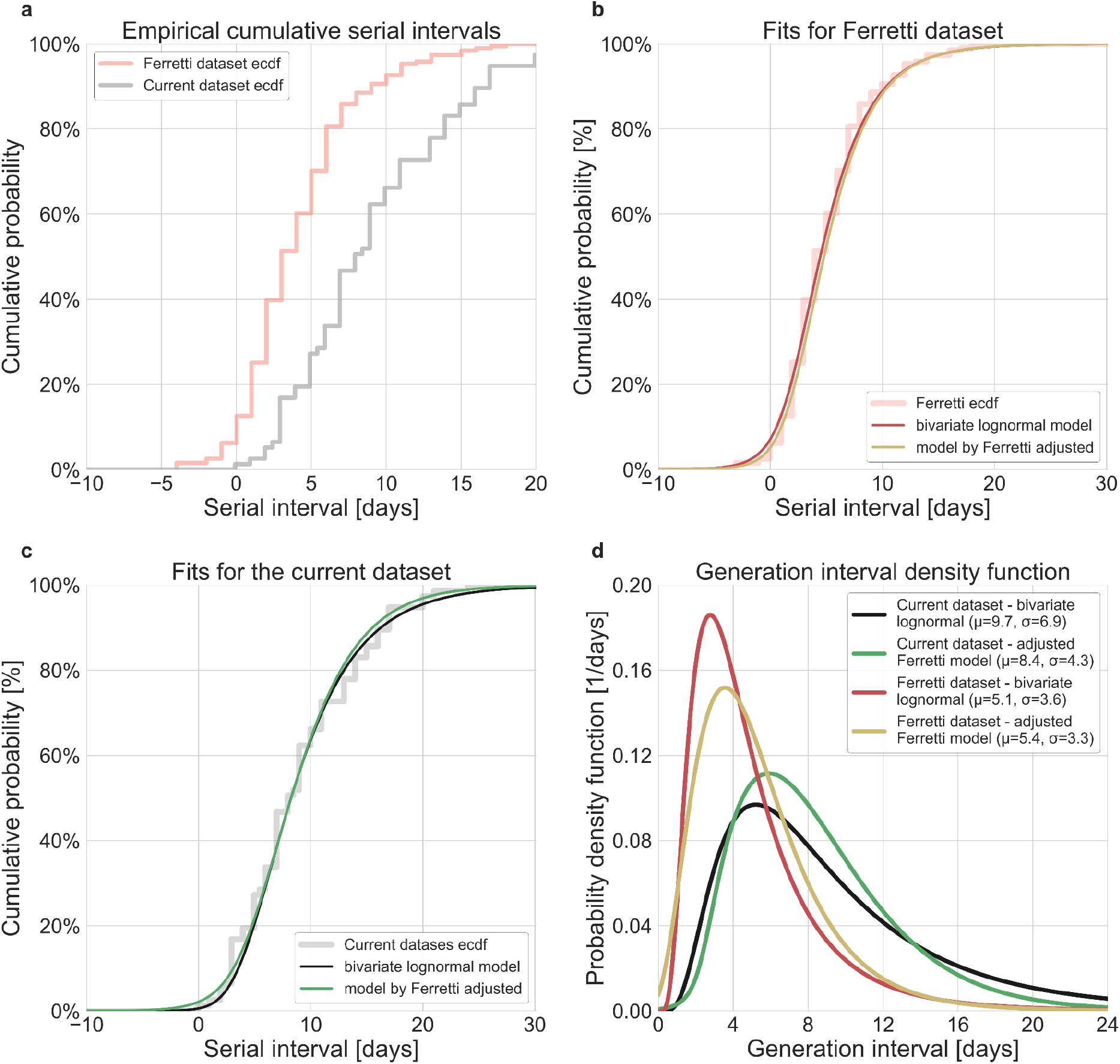
Comparison of the current dataset and model with that of Ferretti et al. The maximum likelihood framework was used to fit both the current dataset and the one provided in the Supplementary Figure S1 of Ferretti et al. The datasets were fit using either the lognormal bivariate model described in the Method section, or a reconstructed model following ^3^ adjusted by adding a parameter for shifting the TOST function over the x-axis. a, The empirical cumulative distribution of serial intervals, comparison between the dataset of Ferretti et al. ^3^ and the current dataset curated in this study. b, Maximum likelihood fits for the dataset provided in the Supplementary Figure S1 of Ferretti et al. c,. Maximum likelihood fits for the current dataset. d,. The marginal generation interval distributions of the maximum likelihood fits. The mean and standard deviation are provided in the legend.

**Figure S10:**
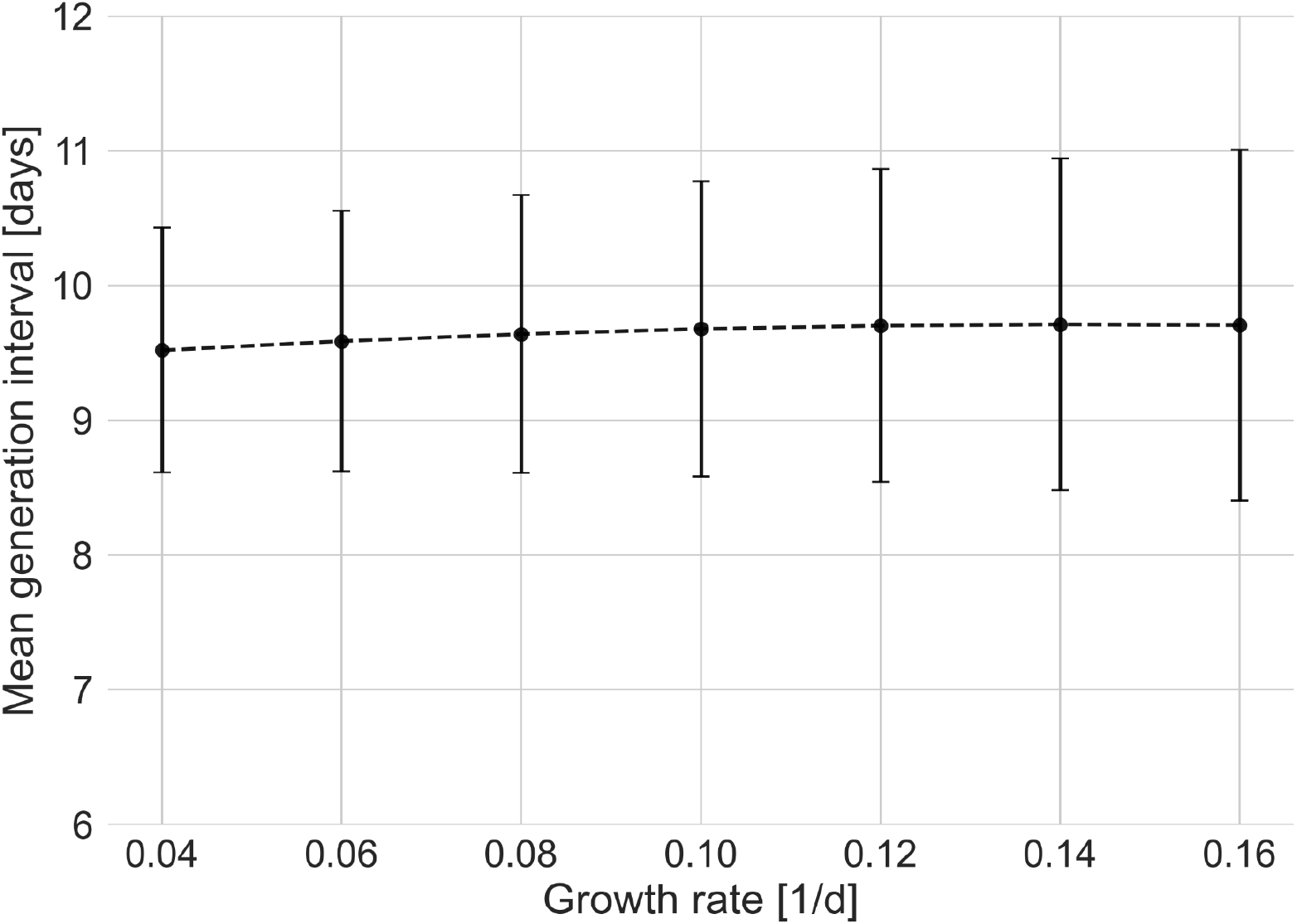
Sensitivity analysis to the growth rate. The mean of the generation interval distributions were estimated using the maximum likelihood fits for the dataset with growth rates in the range of 0.04-0.16 d^-1^. Estimates of the uncertainty were obtained using bootstrapping.

**Figure S11:**
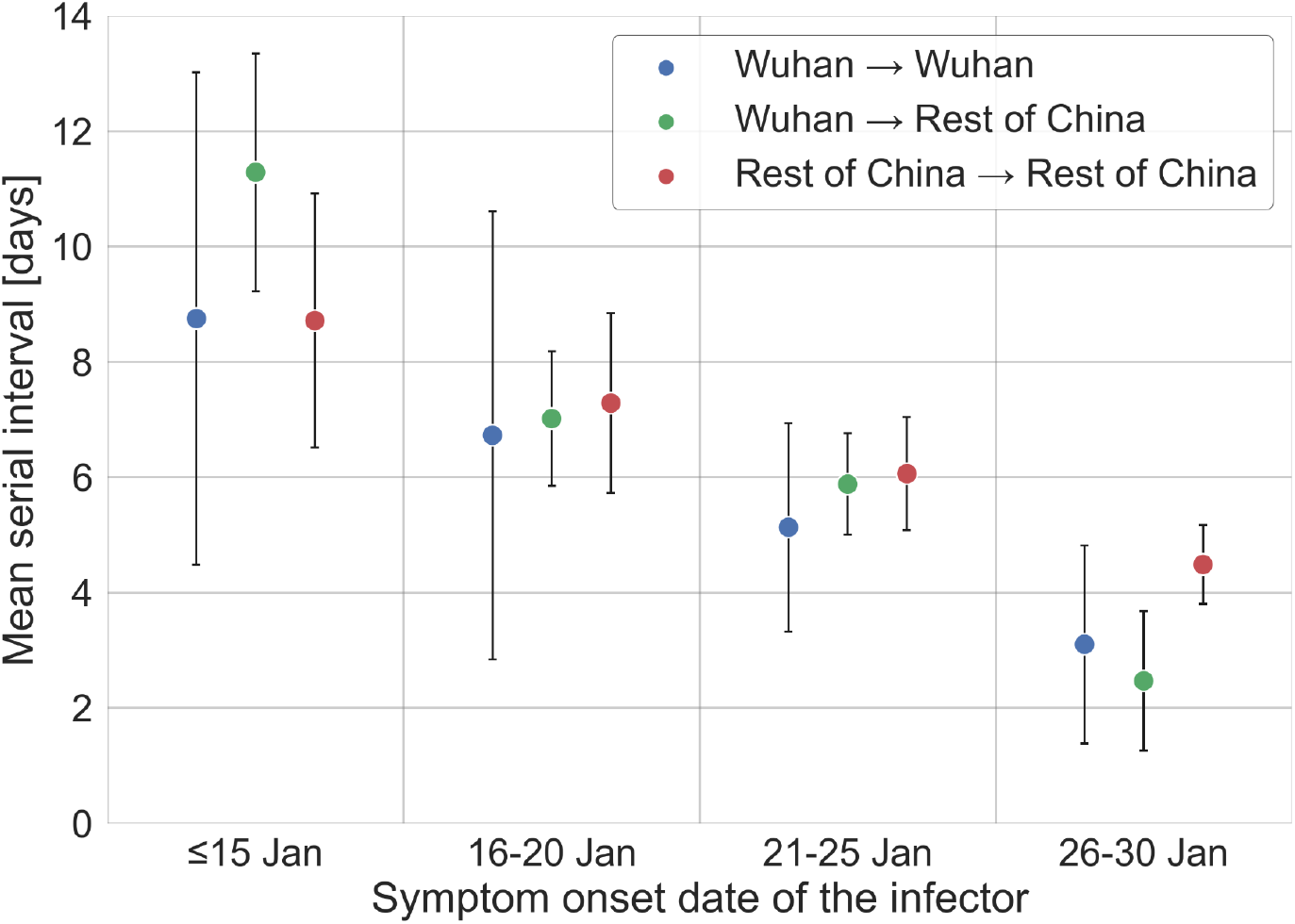
Stratification of the serial interval data by the location of infection. A comparison of the mean of the observed distribution of serial intervals divided to four time periods of the infector symptom onset, and stratified by the infection location of the infector and infectee.

**Figure S12:**
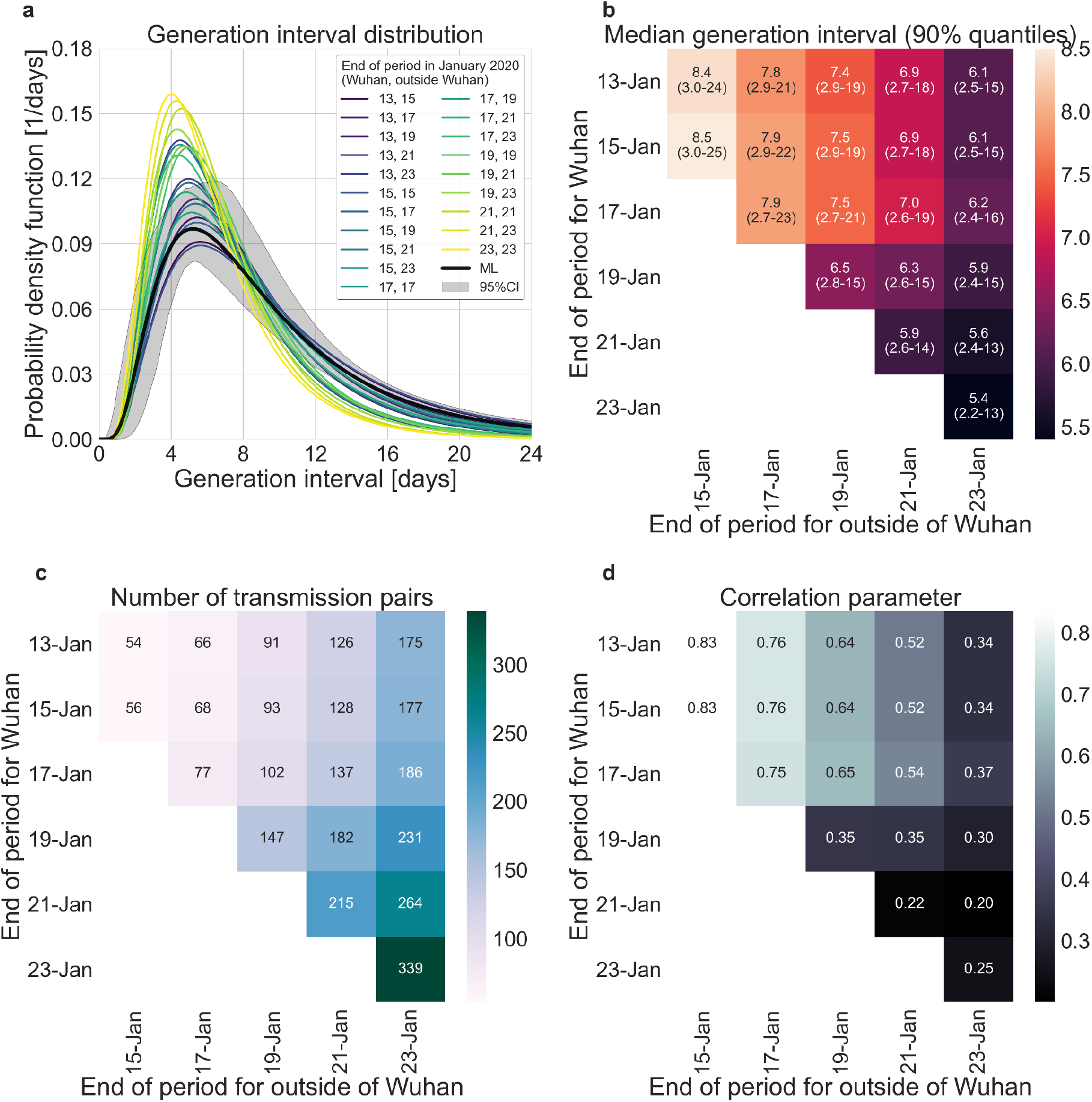
Sensitivity analysis to the definition of the period of interest for infectors that were infected in or outside Wuhan. A comparison of the resulting maximum likelihood estimates where the period of interest was defined separately for infectors who were infected in or outside of Wuhan. **a**, The shapes of the resulting generation interval distribution. For comparison, the main analysis’ maximum likelihood is presented together with its 95% interval (the highlighted area). **b**, Estimates of the median generation intervals and the 90% interquartile-range as function of the period of interest, defined separately for infectors who were infected in or outside of Wuhan. **c**, Number of transmission pairs analysed as function of the period of interest, defined separately for infectors who were infected in or outside of Wuhan. **d**, Estimates for the correlation between incubation period and generation interval parameter as function of the period of interest, defined separately for infectors who were infected in or outside of Wuhan.

**Figure S13:**
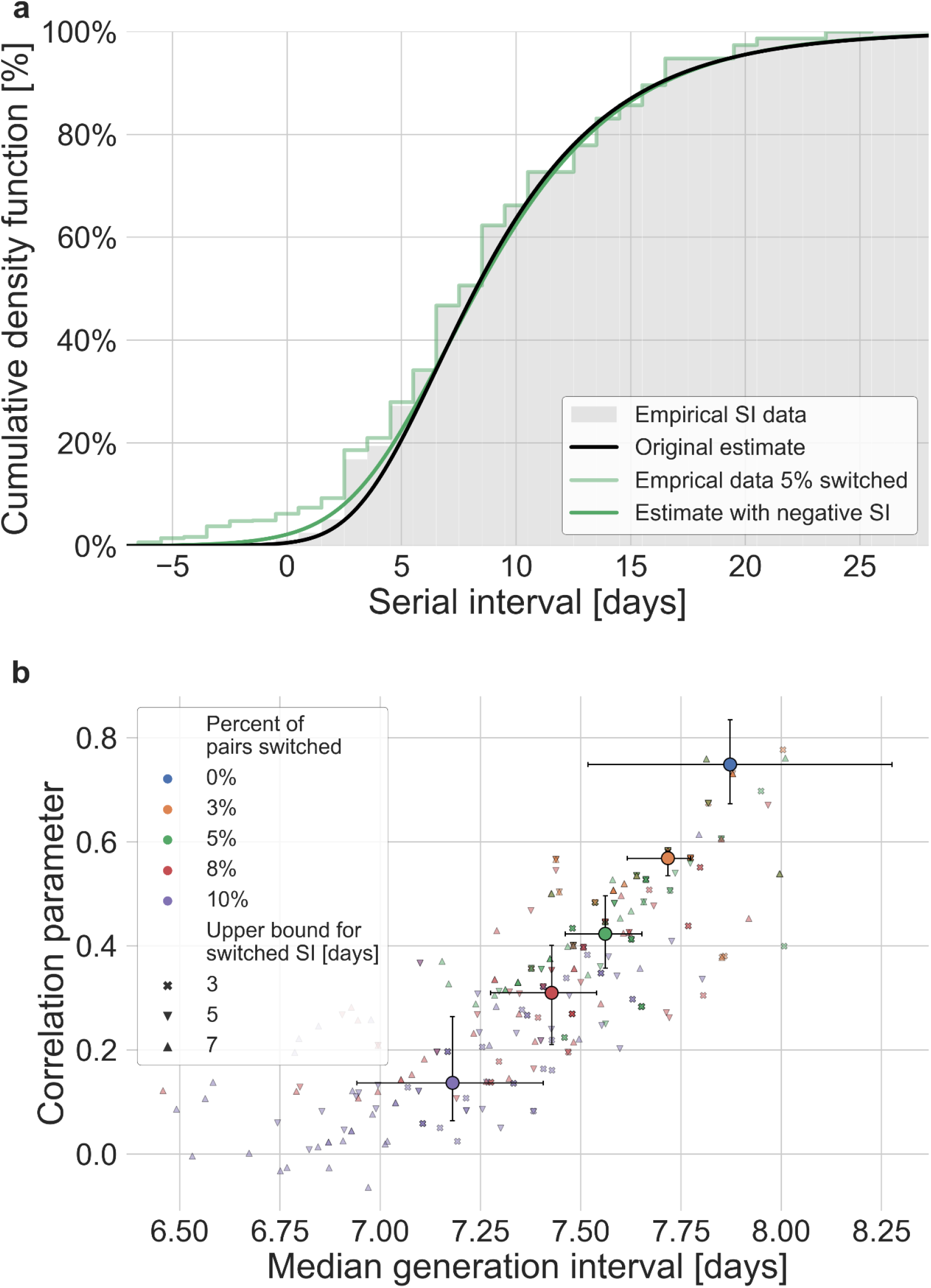
Sensitivity analysis to the pairs’ order of infections. Estimates for the bivariate incubation period and generation interval distribution were obtained for adjusted datasets in which the order of transmission was switched between the infector and infectee (giving a negative serial interval). The analysis was performed by varying the fraction of pairs switched (0-0.1) and the maximal serial interval for which order switching is allowed (3-7 days). For each combination, the analysis was run 30 times while switching the pairs at random. **a**, The serial interval cumulative distribution averaged over all (90) runs with 5% of the pairs switched (chosen as an example). The estimate for the distribution’s parameter was taken as the median across 90 runs. For comparison the original observed serial intervals cumulative distribution and the fit of the models are given. **b**, The resulting estimates of the bivariate incubation period and generation interval distributions are presented via the correlation parameters and median generation intervals. Each point represents a single run, given a percent of pairs switched and a threshold value. The large circles represent the median of the estimates when aggregating runs with a given percent of switched pairs, with error bars corresponding to their interquartile range (25%-75% of the results). For comparison the blue diamond represents the original estimate of the current study, with its uncertainty estimate.

**Figure S14:**
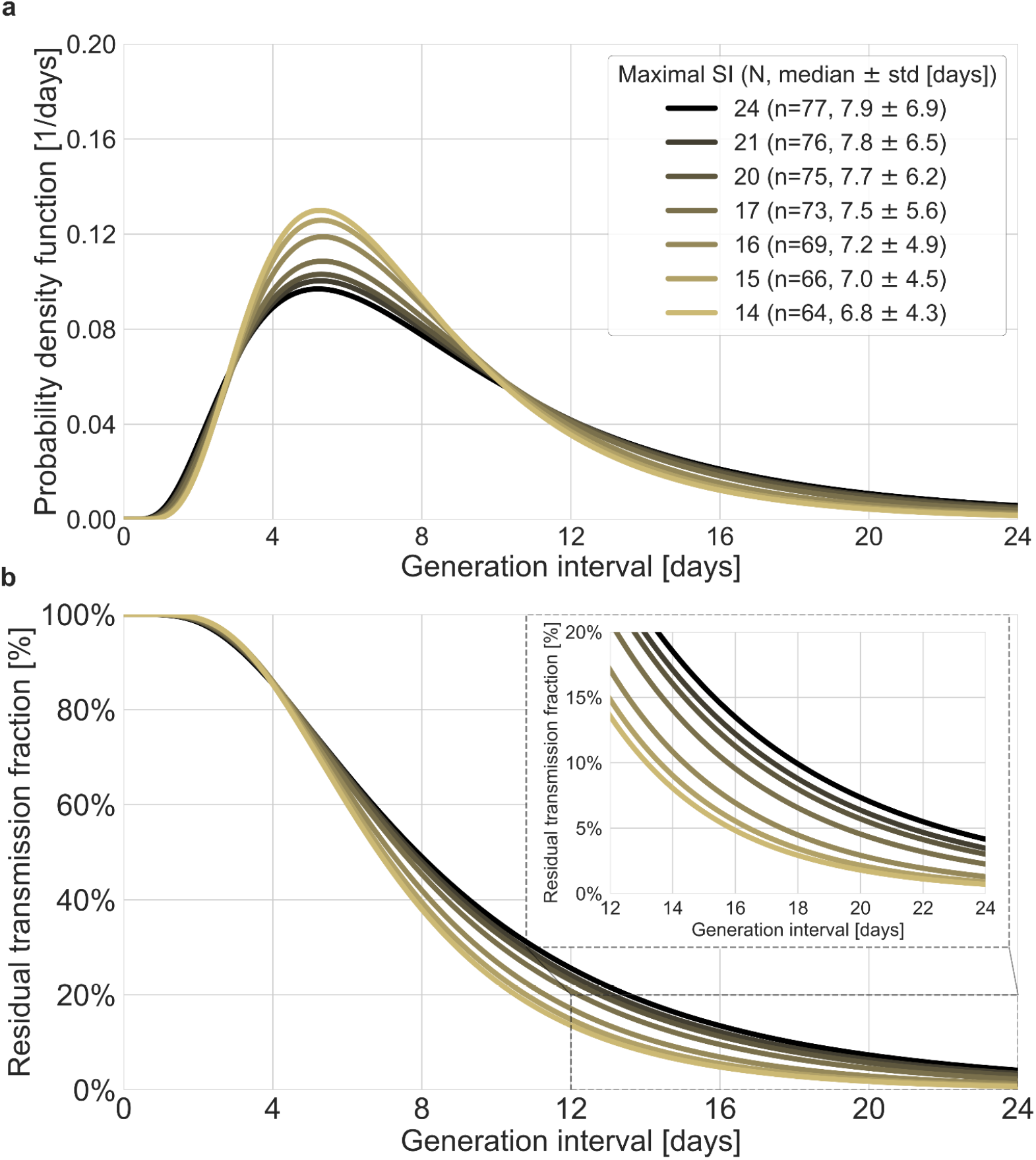
Sensitivity analysis to the top values of serial intervals. The generation-interval distribution is inferred by maximum likelihood when the transmission pairs with the highest serial intervals are removed. **a**, The probability density functions of the distributions. The legend reports the median and standard deviation of each of the distributions, as well as the number of transmission pairs remaining in the dataset after the removal of serial intervals exceeding the specified value. **b**, The survival function of the generation interval distribution, defined as the complement of the cumulative distribution, representing the residual fraction of transmission after a designated time since infection. The inset shows a zoom-in on the period of 10-24 days after exposure, a period in which there is a substantial difference between the current estimate and those from previous studies.

## Supplementary information

### Extended methods

#### Literature survey for serial-interval data

A literature survey was conducted in order to gather data on serial intervals of transmission events from published and preprint literature. The survey was composed using a “google scholar” inquiry containing the phrases: “serial interval” + “COVID” + “china”. Twelve relevant datasets were identified: ^10–12,16–24^

#### Calculation of the mean serial interval for cohorts of transmission pairs that occurred on the same day

In order to compensate for the scarce data with early dates of infector’s onset, we used a simple probabilistic model with Baysian inference to derive crude estimates of the mean serial interval as a function of the infector symptoms onset date. For each date, the serial intervals of infectors that developed symptoms on that day, were assumed to have a Student t distribution such that the mean, standard deviation and degrees of freedoms were random variables sampled from normal, uniform and exponential distributions accordingly. A markov chain monte carlo method was then used to estimate the mean serial interval and its uncertainty (See Results and Figure 2b).

#### Sensitivity analysis to the period of interest

For each of the dates between January 11-25, 2020, we extracted the dataset consisting of the transmission pairs with infector onset symptoms up to that date. We rerun the maximum likelihood framework on the extracted datasets. Furthermore, in order to obtain uncertainty estimates we used a bootstrapping method. In the bootstrapping process we resampled with replacements 100 times and processed via the maximum likelihood framework. In addition, the growth rate (*r*) was sampled from the distribution found by a study of the early outbreak in China ^30^

#### Sensitivity analysis to the infection location of the infector

The transmission pairs dataset contains data from various cities and provinces in China. The mitigation steps were enacted at different time points across China, first in Wuhan and later in other cities and provinces. Previous analysis showed substantial growth rate differences across provinces ^41^, but it seems that when corrected for case ascertainment, the observed difference in growth rate between Wuhan and the rest of China is small (0.08/d vs. 0.1/d) ^30^.

In our main analysis, we do not differentiate transmission pairs by location. Thus, spatial effects could affect our results in two ways: via the estimated growth rate or via the period chosen for analysis as an approximation for unmitigated transmission. Figure S10 shows the sensitivity of the results to a change in the growth rate in the range of 0.04-0.16 d^-1^; Estimates for the mean generation interval change in the range of 8.1-9.1 days. Specifically, assuming a growth rate of 0.08/d instead of 0.1/d has a minimal effect on the main results of the analysis.

We further test how the duration of unmitigated period affects the results of the analysis when the dataset is stratified by the infection location of the infectors and infectee. Figure S11 compares the mean observed serial intervals when the infector and infectee were infected in or outside of Wuhan. We expect that Wuhan to Wuhan transmissions will be shorter than transmissions from Wuhan to the rest of China, but we do not find significant differences, as the data for transmission pairs from Wuhan is scarce. We also check the sensitivity of the generation-interval-distribution estimates to our choices of the unmitigated period, when it is defined separately for those pairs whose infector have been infected inside or outside Wuhan (shown in figure S12). Our analysis suggests that reasonable changes in the unmitigated period have minor effects on our main estimates (e.g. the median generation interval and the 90% of the distribution). For example, taking only pairs with an infector that was infected in Wuhan and developed symptoms until January 15 or pairs with an infector that was infected outside of Wuhan and developed symptoms until January 21 leads to a median generation interval of 6.9 days, in comparison to 7.9 days in our main analysis. Both are substantially larger than previous reports ^3,7,11^.

#### Comparison with another model of infectiousness

To check whether these results are sensitive to our choice of using a bivariate lognormal distribution to characterize the joint distribution of the generation interval and the incubation period, we repeated our analysis using a different functional form using an adjusted logistic TOST model following ^3^. Ferretti et al. modelled the transmission by assuming a TOST distribution with a skewed-logistic shape that is dependent on the incubation period (only on the left side).

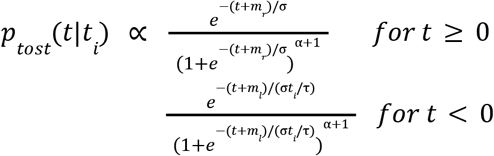

While *t*_*i*_ is the specific incubation period, *τ* is the mean incubation period (5.42 days), *α, σ* are parameters determining the shape of the distribution, and *m*_*1*_, *m*_*r*_ are the median of the distribution of the negative and positive sides 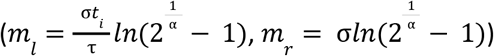.

We adjusted their model by an additional parameter (*l*) enabling shifting the TOST along the time axis.

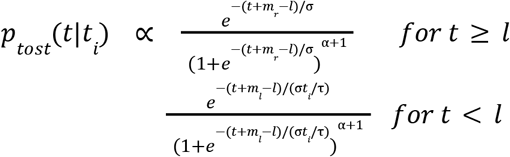

We use our maximum likelihood framework to estimate the generation interval distribution of the adjusted form based on our compiled dataset. Furthermore, we fitted both our models and the adjusted model to the serial interval dataset provided in Ferretti et al. supplementary Figure S1. Results of the comparison are presented in Figure S9.

